# Genome-wide meta-analysis of over 29,000 people with epilepsy reveals 26 loci and subtype-specific genetic architecture

**DOI:** 10.1101/2022.06.08.22276120

**Authors:** International League Against Epilepsy Consortium on Complex Epilepsies, Samuel F Berkovic, Gianpiero L Cavalleri, Bobby PC Koeleman

**Author notes:** **Corresponding authors** Samuel F Berkovic, Gianpiero L Cavalleri, Bobby PC Koeleman. Author names and contributions listed at the end (International League Against Epilepsy Consortium on Complex Epilepsies).

## Abstract

Epilepsy is a highly heritable disorder affecting over 50 million people worldwide, of which about one-third are resistant to current treatments. Here, we report a trans-ethnic GWAS including 29,944 cases, stratified into three broad- and seven sub-types of epilepsy, and 52,538 controls. We identify 26 genome-wide significant loci, 19 of which are specific to genetic generalized epilepsy (GGE). We implicate 29 likely causal genes underlying these 26 loci. SNP-based heritability analyses show that common variants substantially close the missing heritability gap for GGE. Subtype analysis revealed markedly different genetic architectures between focal and generalized epilepsies. Gene-set analysis of GGE signals implicate synaptic processes in both excitatory and inhibitory neurons in the brain. Prioritized candidate genes overlap with monogenic epilepsy genes and with targets of current anti-seizure medications. Finally, we leverage our results to identify alternate drugs with predicted efficacy if repurposed for epilepsy treatment.

## Introduction

The epilepsies are a heterogeneous group of neurological disorders, characterized by an enduring predisposition to generate unprovoked seizures.^1^ It is estimated that over 50 million people worldwide have active epilepsy, with an annual cumulative incidence of 68 per 100,000 persons.^2^

Similar to other common neurodevelopmental disorders, the epilepsies have substantial genetic risk contributions from both common and rare genetic variation. Analysis of the epilepsies benefits from deep phenotyping which allows clinical subtypes to be distinguished^3^, in contrast to other common neurodevelopmental disorders where phenotypic subtypes are more difficult to define. Differences in the genetic architecture of these clinical subtypes of epilepsies are also emerging to complement the clinical partitioning.^4–7^ The rare but severe epileptic encephalopathies are usually non-familial and are largely caused by single *de novo* dominant variants, often involving genes encoding ion channels or proteins of the synaptic machinery.^8^ Common and rare variations have both been shown to contribute to the milder and more common focal and generalized epilepsies. This is particularly true for generalized epilepsy, which is primarily constituted by genetic generalized epilepsy (GGE).^4,5,9,10^ Nevertheless, previous genetic studies of common epilepsies have explained only a few percent of this common genetic variant, or SNP-based, heritability.^4–6,10^

Epilepsy is typically treated using anti-seizure medications (ASMs). However, despite the availability of over 25 licensed ASMs worldwide, a third of people with epilepsy experience continuing seizures.^11^ Diet, surgery and neuromodulation represent additional treatment options that can be effective in small subgroups of patients.^12^ Accurate classification of clinical presentations is an important guiding factor in epilepsy treatment.

Here, we report the third epilepsy GWAS meta-analysis, comprising a total of 29,944 deeply phenotyped cases recruited from tertiary referral centres, and 52,538 controls, approximately doubling the previous sample size.^4^ Results suggest markedly different genetic architectures between focal and generalized forms of epilepsy. Combining these results with results from less stringently phenotyped biobank and deCODE genetics epilepsy cases did not substantially increase signal, despite almost doubling the sample size to 51,678 cases and 1,076,527 controls. Our findings shed light on the enigmatic biology of generalized epilepsy and the importance of accurate syndromic phenotyping, and may facilitate drug repurposing for novel therapeutic approaches.

## Results

### Study overview

We performed a genome-wide meta-analysis by combining the previously published effort from our consortium^4^ with unpublished data from the Epi25 collaborative^10^ and four additional cohorts (**Supplementary table 1**). Our primary mixed model meta-analysis constitutes 4.9 million SNPs tested in 52,538 controls and 29,944 people with epilepsy, of which 16,384 people had neurologist classified focal epilepsy (FE) and 7,407 people had GGE. The epilepsy cases were primarily of European descent (92%), with a smaller proportion of African (3%) and Asian (5%) ancestry (**Supplementary table 2**). Cases were matched with controls of the same ancestry and GWAS were performed separately per ancestry, before performing trans-ethnic meta-analyses for the broad epilepsy phenotypes ‘FE’ (n=16,384 cases) and ‘GGE’ (n=7,407 cases). We further conducted meta-analyses in subjects of European ancestry of the well-defined GGE subtypes of: a) juvenile myoclonic epilepsy (JME), b) childhood absence epilepsy (CAE), c) juvenile absence epilepsy (JAE), and d) generalized tonic-clonic seizures alone (GTCSA), as well as the focal epilepsy subtypes of: a) focal epilepsy with hippocampal sclerosis, b) focal epilepsy with other lesions, and c) lesion-negative focal epilepsy. We ran a variety of follow-up analyses to identify potential sex-specific signals and obtain biological insights and opportunities for drug-repurposing. Sample size prevented inclusion of other ethnicities in the subtype analyses.

### GWAS for the epilepsies

Our ‘all epilepsy’ meta-analysis revealed four genome-wide significant loci, of which two were novel (**Figure 1**). Similar to our previous GWAS^4^, the 2q24.3 locus was composed of two independently significant signals (**Supplementary table 3**). Furthermore, a novel suggestive signal (rs4932477, p=5.04×10^−8^) was found on chromosome 15, containing *POLG*, which is associated with one of the most severe kinds of intractable monogenic epilepsy.^13^ Using ASSET to determine the extent of FE and GGE-related pleiotropy, the 2q24.3 and 9q21.13 signals showed pleiotropic effects at a genome-wide significance level, with concordant SNP effect directions for both forms of epilepsy (**Supplementary table 4**). The 2p16.1 and 10q24.32 loci were primarily derived from GGE. The FE analysis did not reveal any genome-wide significant signals.

**Figure 1.**
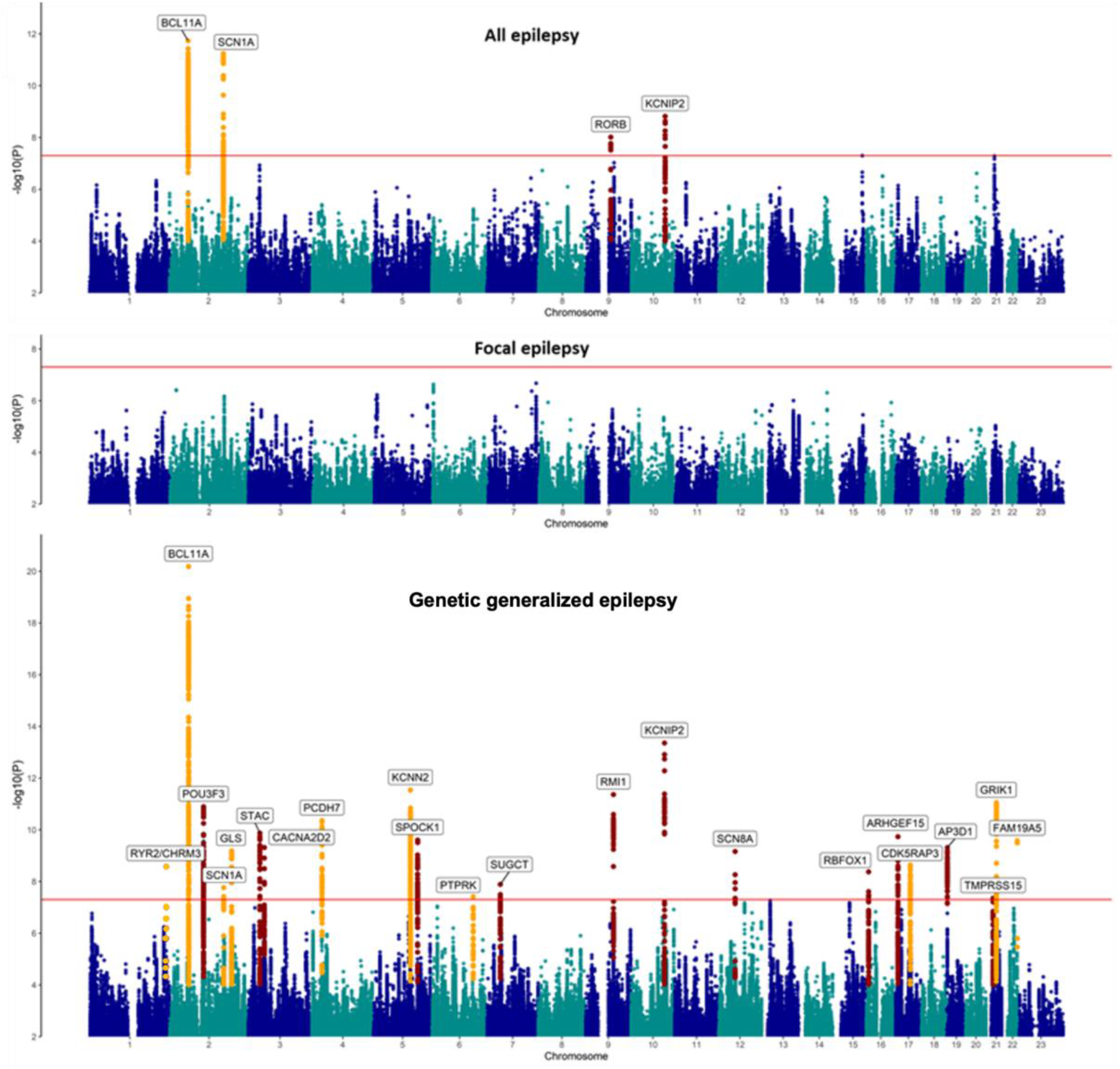
Manhattan plot of trans-ethnic all, focal epilepsy and genetic generalized epilepsy genome-wide meta-analyses. The red line shows the genome-wide significance threshold (5×10^−8^). Chromosome and position are displayed on the x axis and -log10 P-value on the y axis. Novel genome-wide significant loci are highlighted in red and loci previously associated with epilepsy are labelled in orange. Annotated genes are those implicated by our gene prioritization analyses.

Analysis of GGE cases only uncovered a total of 25 independent genome-wide significant signals across 22 loci, of which 13 loci are novel. The strongest signal of association (p=6.6×10^−21^), located at 2p16.1, constitutes three independently significant signals. Similarly, the novel locus 12q13.13 was composed of two independently significant signals (**Supplementary table 3**).

Functional annotation of the 2,355 genome-wide significant SNPs across the 22 GGE loci revealed that most variants were intergenic or intronic (**Supplementary data 1**). 26/2355 (1.1%) SNPs were exonic, of which 12 were located in protein-coding genes and nine were missense variants. Sixty-one percent of SNPs were located in open chromatin regions, as indicated by a minimum chromatin state of 1-7.^14^ Further annotation by Combined Annotation-Dependent Depletion (CADD) scores predicted 110 associating SNPs to be deleterious (CADD score >12.37).^15^ LDAK heritability analyses showed significant enrichment of signal in “super-enhancers” (**Supplementary table 5**), suggesting that GGE variants regulate clusters of transcriptional enhancers that control expression of genes that define cell identity.^16^

To assess potential syndrome-specific loci, we performed GWAS on seven well-defined FE and GGE subtypes **(Supplementary figure 1A-G)**. We found three genome-wide significant loci associated with JME (n=1,813), of which one was novel (8q23.1), and the other two (4p12 and 16p11.2) were reported in our previous GWAS.^4^ All three signals appear specific to JME; without reaching nominal significance in any other GGE subtype. Furthermore, these loci did not reach genome-wide significance when these subtypes were pooled in the GGE analysis. Our analysis of CAE (n=1,072) consolidated an established genome-wide significant signal at 2p16.1, which was also observed in the GGE and all epilepsy GWAS. We did not find any genome-wide significant loci for JAE (n=671), GTCSA (n=499), ‘non-lesional focal epilepsy’ (n=6,367), ‘focal epilepsy with hippocampal sclerosis’ (n=1,375), or ’focal epilepsy with other lesions’ (n=4,661).

Genomic inflation was comparable to our previous GWAS and all linkage-disequilibrium score regression (LDSR) intercepts were lower than in our previous GWAS (**Supplementary table 6**),^4^ suggesting that the signals are primarily driven by polygenicity, rather than by confounding or population stratification.^17^

### Locus annotation, transcriptome-wide association study (TWAS) and gene prioritization

Using FUMA^18^ (see Methods), the ‘all epilepsy’ meta-analysis was mapped to 43 genes and the GGE analysis to 278 genes (**Supplementary data 2**). Thirty nine of the 43 ‘all epilepsy’ genes overlapped with GGE, resulting in a total of 282 uniquely mapped genes. These 282 genes were enriched for monogenic epilepsy genes (hypergeometric test, 18/837 genes overlapped; odds ratio [OR]=1.51, P=0.04), and targets of ASMs (hypergeometric test, 9/191 genes overlap; OR=3.39, P=5.4×10^−4^).

We calculated a gene-based association score based on the aggregate of all SNPs inside each gene using MAGMA (see Methods).^19^ This analysis yielded 39 significant genic associations, six with ‘all epilepsy’, and 37 with GGE (four overlapped with the ‘all epilepsy’ analysis), after correction for 16,371 tested genes (p<0.05/16,371 genes; **Supplementary data 3**). Thirteen of these 39 genes mapped to regions outside of the genome-wide significant loci from the single SNP analyses.

Next, we performed a transcriptome-wide association study (TWAS) to assess whether epilepsy was associated with differential gene expression in the brain (see Methods).^20,21^ These analyses revealed significant associations of 27 genes total; 13 genes with ‘all epilepsy’, 16 with GGE and two with both phenotypes (**Supplementary data 4**). Nineteen of the 27 genes mapped outside of the 26 loci identified through the GWAS. Using Summary-data-based Mendelian Randomization (SMR)^22^, we determined a potentially causal relationship between brain expression of *RMI1* and ‘all epilepsy’, and between *RMI1, CDK5RAP3, TVP23B* and GGE (**Supplementary data 5**).

Of note, expression of *RMI1* was associated with GGE in both TWAS (p=4.0×10^−10^) and SMR (p=5.2×10^−8^), as well as with ‘all epilepsy’ (TWAS p=1.3×10^−6^; SMR p=2.6×10^−6^). *RMI1* has a crucial role in genomic stability^23^ and has not been previously associated with epilepsy nor any other Mendelian trait (OMIM #610404).

We used a combination of ten different criteria to identify the most likely implicated gene within each of the 26 associated loci from the meta-analysis (see Methods). This resulted in a shortlist of 29 genes (**Figure 2**), of which ten are monogenic epilepsy genes, seven are known targets of currently licensed ASDs and 17 are associated with epilepsy for the first time. Interrogation of the Drug Gene Interaction Database (DGIdb) showed that 13 of the 29 genes are targeted by a total of 214 currently licensed drugs (**Supplementary data 6**).

**Figure 2.**
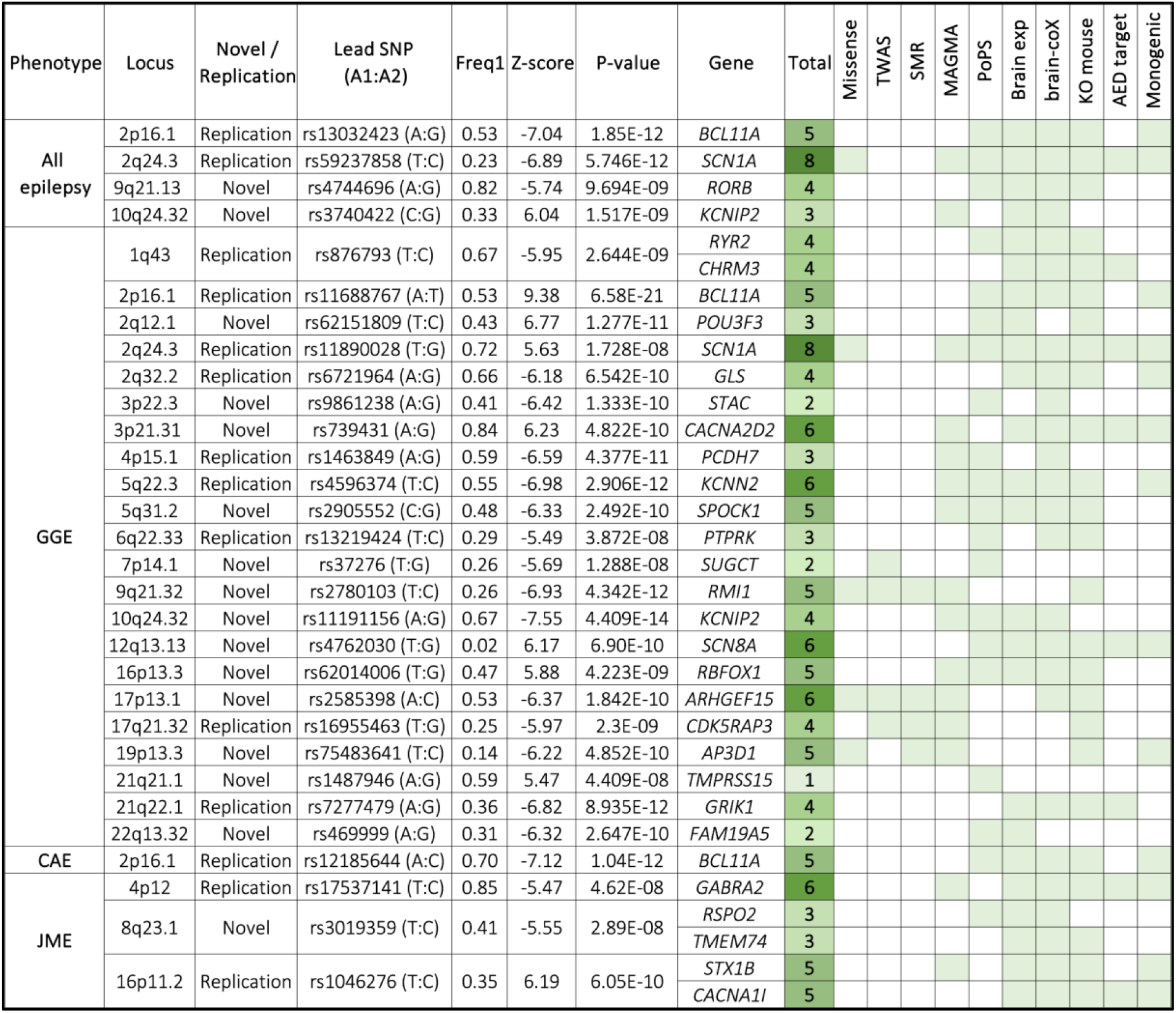
Genome-wide significant loci and prioritized genes. Genome-wide significant loci are annotated with details from the lead-SNP and prioritized genes. Loci were classified as novel or replication according to the genome-wide significant results of previous GWAS publications. Genes were scored based on 10 criteria/methods, after which the gene with the highest score in the locus was selected as the prioritized gene. Total: number of satisfied criteria for gene prioritization. Missense: the locus contains a missense variant in the gene. TWAS: significant transcriptome-wide association with the gene. SMR: significant summary-based mendelian randomisation association with the gene. MAGMA: significant genome-wide gene based association. PoPS: gene prioritized by polygenic priority score. Brain exp: the gene is preferentially expressed in brain tissue. Brain-coX: the gene is prioritized as co-expressed with established epilepsy genes. KO mouse: knockout of the gene causes a neurological phenotype in mouse models. Monogenic: the gene is a known cause of monogenic epilepsy. Genomic coordinates for each locus (hg19) can be found in Supplementary table 3.

The strongest association signal for GGE was found at 2p16.1, consistent with our previous results where we implicated the gene *VRK2* or *FANCL*.^24^ Our gene prioritization analysis now points to the transcription factor *BCL11A* as the culprit gene, located 2.5MB upstream of the lead SNPs at this locus. Two of three lead SNPs are located in enhancer regions (as assessed by chromatin states in brain tissue) which are linked to the *BCL11A* promoter via 3D chromatin interactions (**Supplementary figure 2**). Rare variants in *BCL11A* were recently associated with intellectual disability and epileptic encephalopathy.^25^ However, interrogation of the MetaBrain eQTL database did not reveal a significant association of our lead SNPs with *BCL11A* expression.

### The HLA system and common epilepsies

The highly polymorphic HLA region has been associated with various neuropsychiatric and autoimmune neurological disorders following accurate capture of all its genetic variation. Therefore, we imputed HLA alleles and amino acid residues using CookHLA^26^ and ran association across epilepsy, focal and GGE phenotypes, as well as the seven sub-phenotypes (see Methods). No SNP, amino acid residue or HLA allele reached the level of genome-wide significance (see **Supplementary figure 3**). The most significant signal was with GGE, in which an aspartame amino acid residue in exon 2 position 31432494 had a p-value of 3.8×10^−7^.

**Figure 3.**
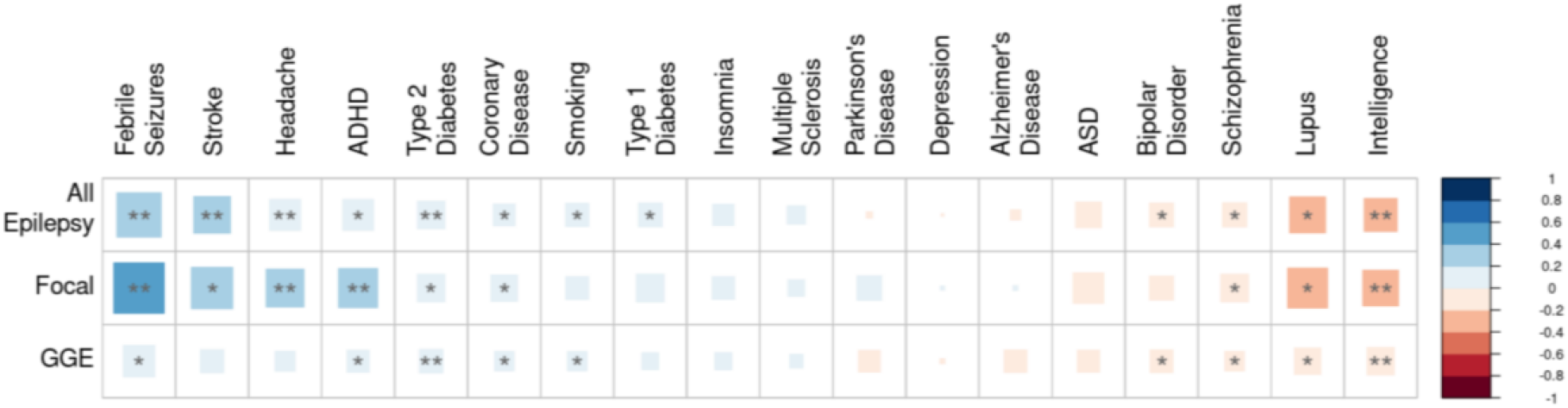
Genetic correlations of epilepsy with other phenotypes. The genetic correlation coefficient was calculated with LDSC and is denoted by color scale from -1 (red; negatively (anti-)correlated) to +1 (blue; positively correlated). The square size relates to the absolute value of the corresponding correlation coefficient.* P < 0.05, ** P < 0.0009 (Bonferroni corrected).

### SNP-based heritability

We calculated SNP-based heritability using LDAK to determine the proportion of epilepsy risk attributable to common genetic variants. We observed liability scale SNP-based heritabilities of 17.7% (95% CI 15.5 - 19.9%) for all epilepsy, 16.0% (14.0 - 18.0%) for FE and 39.6% (34.3 - 44.6%) for GGE. Heritabilities for GGE subtypes were notably higher for all individual GGE subtypes: ranging from 49.6% (14.0% - 85.3%) for GTCSA to 90.0% (63.3 - 116.6%) for JAE (**Supplementary table 7**).

Employing a univariate causal mixture model^27^ (see Methods) we estimated that 2,850 causal SNPs (standard error: 200) underlie 90% of the SNP-based heritability of GGE, comparable with previous estimates.^9^ Power analysis demonstrated that the current genome-wide significant SNPs only explain 1.5% of the phenotypic variance, whereas an estimated sample size of around 2.5 million subjects would be necessary to identify the causal SNPs that explain 90% of GGE SNP-based heritability (**Supplementary figure 4)**.

To further explore the heritability of the different epilepsy phenotypes, we used LDSC to perform genetic correlation analyses.^28^ We found evidence for strong genetic correlation between all four GGE syndromes (**Supplementary figure 5**). We also observed a significant genetic correlation between the focal non-lesional and JME syndromes, which has been reported previously.^4^ Here, with larger sample sizes, CAE also showed a significant genetic correlation with the focal non-lesional cohort.

### Tissue and cell-type enrichment

To further illuminate the underlying biological causes of the epilepsies, we used MAGMA^19^ and data from the gene-tissue expression consortium (GTEx) to assess whether our GGE-associated genes were enriched for expression in specific tissues and cell types (see Methods). We identified significant enrichment of associated genes expressed in brain and pituitary tissue (**Supplementary figure 6**). This is the first time the pituitary gland has been implicated in GGE and might reflect a hormonal component to seizure susceptibility. Further sub-analyses showed that our results were enriched for genes expressed in almost all brain regions, including subcortical structures such as the hypothalamus, hippocampus and amygdala (**Supplementary figure 7**). We did not find enrichment for genes expressed at specific developmental stages in the brain (**Supplementary figure 8**).

Cell-type specificity analyses of GGE data using various single-cell RNA-sequencing reference datasets (see Methods) revealed enrichment in excitatory as well as inhibitory neurons, but not in other brain cells like astrocytes, oligodendrocytes or microglia (**Supplementary figure 9**). Similarly, stratified LD-score regression using single-cell expression data (see Methods) did not reveal a difference between excitatory and inhibitory neurons (p=0.18).

### Gene-set analyses

MAGMA gene-set analyses showed significant associations between GGE and biological processes involving various functions in the synapse (**Supplementary data 7**). To further refine the synaptic signal, we performed a gene-set analysis using lists of expert-curated gene-sets involving 18 different synaptic functions.^29^ These analyses showed that GGE was associated with intracellular signal transduction (n=139 genes, p=9.6×10^−5^) and excitability in the synapse (n= 54 genes, p=0.0074). None of the other 16 synaptic functions showed any association (**Supplementary data 7**). Genes involved with excitability include the N-type calcium channel gene *CACNA2D2*, implicated at the novel GGE locus 3p21.31. N-type calcium channel blockers such as levetiracetam and lamotrigine are amongst the most widely used and effective ASMs for GGE as well as focal epilepsy.^30–32^ Together, these results suggest that the genes associated with GGE are expressed in excitatory as well as inhibitory neurons in various brain regions, where they affect excitability and intracellular signal transduction at the synapse.

### Sex-specific analyses

There are known sex-related patterns in the epidemiology of epilepsy. Although females have a marginally lower incidence of epilepsy than males, GGE is known to occur more frequently in females.^33^ To test whether this sex divergence has a genetic basis, we performed sex-specific GWAS for all, GGE and FE (**Supplementary figures 10-12**). Analyses revealed one female-specific genome-wide significant signal at 10q24.32 (lead SNP: rs72845653), containing *KCNIP2*, implicated in our main GGE meta-analysis (lead SNP: rs11191156). However, the lead SNPs of these two signals are not in LD (r^2^=0.05). Interestingly, the direction of effect of this signal is opposite in females and males. This sex difference is further corroborated by significant sex-heterogeneity (p=1.54×10^−8^) and gender-differentiation (p=5.6×10^−9^).^34^ Sex-related differences in transcription levels in human heart have previously been reported for *KCNIP2*.^35^ We did not find any sex-divergent signals for ‘all’ or focal epilepsy.

LDSC was used to assess the genetic correlation between male-only and female-only GWAS. The male and female GWAS of all epilepsy, FE and GGE were strongly genetically correlated (all Rg>0.9) and none of these correlations were significantly different from 1 (all p>0.05). These results suggest that, with the exception of the female-specific 10q24.32 signal, the overall genetic basis of common epilepsy appears largely similar between males and females.

### Genetic overlap between epilepsy and other phenotypes

To explore the genetic overlap of epilepsy with other diseases, we first cross-referenced the 26 genome wide epilepsy loci with other traits with significant associations (p<5×10^−8^) for the same SNP, or SNPs in strong linkage disequilibrium with our lead SNPs (as detailed in **figure 2**). This analysis revealed eighteen likely pleiotropic loci, with previous associations reported across a variety of traits, the most common being cognitive, sleep, psychiatric, coronary and blood cell traits (**Supplementary figure 13**). The remaining eight loci appear to be specific to epilepsy (3p22.3, 4p12, 5q31.2, 7p14.1, 8q23.1, 9q21.13, 21q21.1, 21q22.1).

We then performed genetic correlation analyses between 18 selected traits and all, GGE and focal epilepsy using LDSC^17^. The selected traits had either, or a combination of 1) epilepsy as a common comorbidity or 2) pleiotropic loci shared with epilepsy. Significant correlations (P<0.05/54=0.0009) were found with febrile seizures, stroke, headache, ADHD, type 2 diabetes and intelligence (**Figure 3**).

Genetic correlation analyses assess the aggregate of shared genetic variants associated with two phenotypes. However, genetic correlations can become close to zero when there is consistent mixed directionality of SNP effects between two phenotypes.^36^ Autism spectrum disorder (ASD) was not significantly correlated, despite monogenic pleiotropy with epilepsy genes supporting an overlap. To explore whether inverse directionality could explain the lack of genetic correlation between ASD and epilepsy we applied the MiXeR tool to GGE, intelligence and ASD, to quantify polygenic overlap irrespective of genetic correlation (see Methods). Results showed that >99% of causal SNPs underlying GGE are shared with intelligence, of which 58% have a discordant direction of effect (**Supplementary figure 14**). Furthermore, despite a lack of genetic correlation with ASD (Rg=-0.12, p=0.06, all epilepsy; Rg=-0.17, p=0.06 focal epilepsy; Rg=-0.09, p=0.09, GGE), we found that 95% of causal SNPs underlying GGE are shared with ASD, but 59% have a discordant direction of effect. This is consistent with the finding that epilepsy and ASD can have a shared genetic cause.^37,38^ For example, monogenic ASD and epilepsy can occur as the result of pathogenic variants in *SCN2A*. Functional studies have shown that ASD without seizures can be caused by loss-of-function variants in *SCN2A* ^39^, whereas epilepsy can be caused by gain-of-function variants in *SCN2A*.^40,41^ Indeed, ASD variants in *SCN2A* seem protective against neuronal hyperexcitability.^41^

### Leveraging GWAS for drug repurposing

To test the potential of our meta-analysis to inform drug repurposing, we predicted the relative efficacy of drugs for epilepsy (see Methods). This analysis was based upon the predicted ability of each drug to modulate epilepsy-related changes in the function and abundance of proteins, as inferred from the GWAS summary statistics (see Methods).^42^ We validated the drug predictions by determining if they are concordant with findings from clinical experience and trials. In our predictions for all epilepsy, current ASMs were ranked higher than expected by chance (p < 1×10^−6^), and higher than drugs used to treat any other human disease. For GGE, broad-spectrum ASMs were predicted to be more effective than narrow-spectrum antiseizure drugs (p < 1×10^−6^), consistent with clinical experience.^43^ Furthermore, the predicted order of efficacy for GGE of individual ASMs matched their observed order in the largest head-to-head randomized controlled clinical trials for generalized epilepsy,^32,44^ an observation unlikely to occur by chance (p < 1×10^−6^).

Using this approach, we highlight the top 20 drugs that are licensed for conditions other than epilepsy, but are predicted to be efficacious for generalized epilepsy, and additionally have published evidence of antiseizure efficacy from multiple published studies and multiple animal models (**Supplementary table 8**). The full list of all predictions can be found in **Supplementary data 8**.

### GWAS in epilepsies ascertained from population biobanks and from deCODE genetics

We performed GWAS using data from several large-scale population biobanks and from deCODE genetics (total cases n=21,734, total controls n=1,023,989, phenotyped using ICD codes, see Methods). Although the biobank and deCODE genetics-specific GWAS did not identify any genome-wide significant loci for GGE or ‘all epilepsy’, one significant locus at 2q22.1 (nearest gene, *NXPH2*) emerged for focal epilepsy (**Supplementary figure 15**).

Meta-analysis of the biobank and deCODE genetics summary statistics with those from the primary epilepsy GWAS identified seven significant loci for the ‘all epilepsy’ phenotype. Six of these signals were previously identified in the primary ‘all epilepsy’ (n=4) or the ‘GGE’ GWAS (n=2). One locus (2q12.1) was novel. The combined biobank and deCODE genetics meta-analysis for GGE identified five novel loci, but four loci from our primary GWAS fell below significance (**Supplementary figure 16**). The combined focal epilepsy meta-analysis showed no significant associations. LDSC between the biobank/deCODE genetics and the primary GWAS results showed genetic correlations ranging between 0.31 and 0.74 (**Supplementary table 9**).

## Discussion

In this study, we leveraged a substantial increase in sample size to uncover 26 common epilepsy risk loci, of which 16 have not been reported previously. Using a combination of ten post GWAS analysis methods, we pinpointed 29 genes that most likely underlie these signals of association. These signals showed enrichment throughout the brain and indicate an important role for synapse biology in excitatory as well as inhibitory neurons. Drug prioritization from the genetic data highlighted licensed ASMs, ranked the ASMs broadly in line with clinical experience and pointed to drugs for potential repurposing. These findings further our understanding of the pathophysiology of common epilepsies and provide new leads for therapeutics.

The 26 associated loci included some notable monogenic epilepsy genes. These include the calcium channel gene *CACNA2D2*, an established epileptic encephalopathy gene^45^ that is directly targeted by ten currently licenced drugs, including two ASMs (gabapentin and pregabalin) as well as the Parkinson’s disease drug safinamide and the nonsteroidal anti-inflammatory drug celecoxib. Both safinamide and celecoxib have evidence of anti-seizure activity.^46,47^ *SCN8A*, which encodes a voltage-gated sodium channel, is an established epileptic encephalopathy gene and is associated with common epilepsies for the first time here. Na_v_1.6 (encoded by *SCN8A)* is targeted by commonly used sodium channel blocking drugs that have been found to be the most efficacious ASMs for people with monogenic *SCN8A-*related epilepsies that are often due to channel gain-of-function.^48^ Additional drugs targeting Na_v_1.6 include safinamide and quinidine. *RYR2* encodes a ryanodine receptor, is an established cardiac disorder gene, has recently been implicated in epilepsy ^49,50^ and is targeted by caffeine as well simvastatin, atorvastatin and carvedilol. The acetylcholine receptor gene *CHRM3* has been previously associated with epilepsy^51^ and is targeted by drugs including solifenacin, used to treat urinary incontinence.

We found that GGE in particular has a strong contribution from common genetic variation. When analyzing individual GGE syndromes, we found that up to 90% of liability is attributable to common variants in the JAE subtype, making it amongst the highest of over 700 traits reported in a large GWAS atlas^52^ (albeit with relatively large confidence intervals; **Supplementary table 7**). The heritability estimates decrease to 40% for the collective GGE phenotype, possibly due to increased heterogeneity from combining syndromes with pleiotropic as well as syndrome-specific risk loci. Although statistical power drastically decreased when assessing specific GGE syndromes, three loci appeared specific to JME. These findings highlight the unique genetic architecture of the subtypes of common epilepsies, which are characterized by a high degree of both shared, and syndrome-specific, genetic risk.

In contrast to GGE, for focal epilepsies we found only a minor contribution of common variants, with no variant reaching genome-wide significance. It would seem that focal epilepsies, as a group, are far more heterogeneous than GGE. Our attempt to mitigate this heterogeneity by performing subtype analysis contrasted with the results from GGE, suggesting different genetic architectures, consistent with the experience from studies of common^9^ and rare^5^ genetic variation and PRS.^6^ There is also emerging evidence for a significant role of non-inherited, somatic mutations in focal epilepsies^.53^

This work highlights the challenges of working with epilepsy cohorts ascertained through large biobanking initiatives. Accurate classification of epilepsy requires a combination of clinical features, electrophysiology and neuroimaging. These details were not available from the biobanks we worked with. Rather, phenotypes were generally limited to ICD codes, which are prone to misclassification.^54^ Population biobanks are also probably ascertaining milder epilepsies that are responsive to treatment, contrasting with the enrichment for refractory epilepsies at tertiary referral centres.

Moreover, a proportion of adults with epilepsy have an acquired brain lesion, such as stroke, tumors or head trauma. Biobanks typically provide self-reported clinical information and codes from primary care and inpatient hospital care episodes, but not neurological specialist outpatient records that would indicate whether previous brain insults were considered relevant to the epilepsy. As a result, the inclusion of the biobank data appeared to introduce more heterogeneity. This contrasts with genetic mapping of other polygenic diseases like type 2 diabetes and migraine, which are relatively easy and reliable to diagnose and classify, resulting in a great increase in GWAS loci when including data from the same biobanks as included in our study.^55,56^

We found enrichment of GGE variants in brain-expressed genes, involving excitatory and inhibitory neurons, but not any other brain cell type. This contrasts with other neurological diseases. For example, microglia are involved in Alzheimer’s disease^57^ and multiple sclerosis,^58^ whereas migraine does not appear to have brain cell specificity.^56^ We further refine this signal by showing an involvement of synapse biology, primarily intracellular signal transduction and synapse excitability. These findings suggest an important role of synaptic processes in excitatory and inhibitory neurons throughout the brain, which could be a potential therapeutic target. Indeed, synaptic vesicle transport is a known target of the ASMs levetiracetam and brivaracetam.^59^

We confirmed that our GWAS-identified genes had significant overlap with monogenic epilepsy genes. A similar convergence of common and rare variant associations has been observed for other neurological neuropsychiatric conditions including schizophrenia^60^ and ALS^61^. The genes prioritized in our GWAS signals also overlapped with known targets of current ASMs^4^ and we have provided a list of other drugs that directly target these genes. Moreover, using a systems-based approach^42^ we highlight drugs that are predicted to be efficacious when repurposed for epilepsy, based on their ability to perturb function and abundance in gene expression. Insights from GWAS of epilepsy have the potential to accelerate the development of new treatments via the identification of promising drug repurposing candidates for clinical trials.^62^ We anticipate that follow-up studies of the highlighted drugs in this study could show clinical efficacy in epilepsy treatment.

In summary, these new data reveal markedly different genetic architectures between the milder and more common focal and generalized epilepsies, provide novel biological insights to disease aetiology and highlight drugs with predicted efficacy when repurposed for epilepsy treatment.

## Methods

### Ethics statement

Local institutional review boards approved study protocols at each contributing site. All study participants provided written, informed consent for use of their data in genetic studies of epilepsy. For minors, written informed consent was obtained from their parents or legal guardian.

### Sample and phenotype descriptions

This meta-analysis combines previously published datasets with novel genotyped cohorts. Descriptions of the 24 cohorts included in our previous analysis can be found in the Supplementary table 6 of that publication.^4^ Here we included 5 novel cohorts (**Supplementary table 1**), comprising 14,732 epilepsy cases and 22,362 controls, resulting in a total sample size of 29,944 cases and 52,538 controls. Classification of epilepsy was performed as described previously.^4^ In brief, we assigned people with epilepsy into focal epilepsy, genetic generalized epilepsy (GGE) or unclassified epilepsy. ‘All epilepsy’ was the combination of GGE, focal and unclassified epilepsy. Where possible, we used EEG, MRI and clinical history to further refine the subphenotypes: juvenile myoclonic epilepsy (JME), childhood absence epilepsy (CAE), juvenile absence epilepsy (JAE), generalized tonic-clonic seizures alone (GTCSA), non-lesional focal epilepsy, focal epilepsy with hippocampal sclerosis (HS) and focal epilepsy with lesion other than HS.

### Genotyping, quality control and imputation

Subjects were genotyped on single nucleotide polymorphism (SNP) arrays, see **Supplementary table 1** for an overview of genotyping in novel cohorts. Quality control (QC) was performed separately for each cohort. Prior to imputation, data from the Janssen, Austrian, Swiss, Norwegian, and BPCCC cohorts were cross-referenced to the HRC panel to ensure SNPs matched in terms of strand, position, and ref/alt allele assignment. Additionally, SNPs were removed if they were absent in the HRC panel, if they had a >20% allele frequency difference with the HRC panel, or if any AT/GC SNPs had MAFs>40%, using tools available from https://www.well.ox.ac.uk/~wrayner/tools/. Data were then imputed using the the Wellcome Sanger Institutes’ imputation server (https://imputation.sanger.ac.uk/), using EAGLE v2.4.1^63^ for phasing, and the Positional Burrows Wheeler Transform algorithm^64^ for imputation. The Haplotype Reference Consortium (HRC) reference panel r1.1 was used as a reference for imputation^65^. Post-imputation, SNPs with an INFO score of <0.9 were removed. The high-INFO SNPs were then converted back to PLINK format and once-again QC’d for genotype coverage (>0.98), minor allele frequencies (>5%) and Hardy-Weinberg Equilibrium violations (p>10^−5^), following previously described methodologies^4^. We removed variants <5% MAF in these 5 cohorts for QC reasons, and note there will be a corresponding loss in study power for the ‘focal’ and ‘all epilepsy’ epilepsy analysis.

QC for the Epi25 cohort was performed using a similar in-house pipeline. Samples were split by ethnicity based on principal component analysis. Pre-imputation QC included filtering of SNPs with call rate (<98%), differential missing rate, duplicated and monomorphic SNPs, SNPs with batch association (p<10^−4^), violation of Hardy-Weinberg Equilibrium (p<10^−10^). Sample filtering included removal of outliers (>4 SD from mean) of heterozygous/homozygous ratio, removal of one of each pair of related samples (proportion identity-by-descent >0.2) and removal of samples with ambiguous or non-matching genetically imputed sex. Furthermore, 3,180 duplicates between the Epi25 cohort and the previously published genome-wide mega-analysis were identified based on genotype, and were removed from the Epi25 cohort. Of the 3,180 duplicates, 1226 were GGE and 1402 focal epilepsy. Genotypes were imputed on the Michigan imputation server, using the Haplotype Reference Consortium v1.1 (n=32470) reference panel for subjects of European and Asian ancestry, and the 1000 Genomes Phase 3 v5 (n=2504) reference panel for subjects of African ancestry. Default imputation parameters and pre-imputation checks were used. Imputed dosages were used for subsequent analyses, filtering on imputation INFO>0.3 and minor-allele frequency >1%.

### Genome-wide association analyses

GWAS of the Janssen Pharmaceuticals, Swiss GenEpa, Norwegian GenEpa and Austrian GenEpa cohorts was performed as a mega-analysis, as described previously.^4^ GWAS of the Epi25 cohort was performed with a generalized mixed model using SAIGE v0.38.^66^ SAIGE was performed in two steps: (1) fitting the null logistic mixed model to estimate the variance component and other model parameters; (2) testing for the association between each genetic variant and phenotypes by applying SPA to the score test statistics. For step 1, SNPs were filtered on call rate >0.98 and MAF >5%, and SNPs were pruned to obtain approximate independent markers (window size of 100 kb and R^2^>0.3), while including sex and the top 10 principal components as covariates. Next, we performed P-value based fixed-effects meta-analyses with METAL^67^ for each of the main phenotypes (all, GGE, and focal epilepsy), as well as the subphenotypes, weighted by effective samples sizes (N_eff_ = 4/(1/N_cases_ + 1/N_controls_)) to account for case-control imbalance. We performed trans-ethnic and European-only meta-analyses for the main phenotypes, and restricted the subphenotype analyses to Europeans only, due to limited sample size in other ethnicities. We included all SNPs (∼4.9 million, MAF>1%) that were present in at least the previous mega-analysis and the Epi25 dataset, which together account for 88% of the total sample size. We calculated genomic inflation factors (λ), mean χ^2^ and LD score regression intercepts to assess potential inflation of the test statistic. Since λ Is known to scale with sample size, we also calculated λ1000, which is λ corrected for an equivalent sample size of 1000 cases and 1000 controls.^68^ We limited these analyses to subjects of European ancestry, since LD-structure depends on ethnicity and Europeans constituted 92% of cases.

### Data sources for the Biobank and deCODE genetics GWAS

Summary statistics for epilepsy GWAS were obtained from three population biobanks; UK Biobank,^69^ Biobank Japan,^70,71^ Finngen release R6,^72^ and from deCODE genetics^73^ (Iceland). The biobank Japan, Finngen and deCODE genetics epilepsy cases were further assigned into either ‘focal’ or ‘generalized’ epilepsy (see below), whereas the UK Biobank samples were not subdivided based on seizure localisation, as the relevant clinical details were unavailable to facilitate an accurate subdivision (see **Supplementary table 10** for sample sizes per biobank and deCODE genetics). Control data were population matched samples with no history of epilepsy.

Fixed-effects meta-analyses were conducted using METAL^67^, weighted by effective sample size (N_eff_ = 4/(1/N_cases_ + 1/N_controls_)) to account for case-control imbalance.

#### UK Biobank

We identified people with epilepsy from the UK Biobank using an analysis of self-reported data, inpatient hospital episode statistics (HES), death certificate diagnostic data and primary care diagnostic data as described elsewhere.^74^ This allowed us to interrogate the evidence available to support a diagnosis of epilepsy rather than relying purely on UK Biobank generated data fields 131048 and 13049 based on ICD-10 G40 mapping.

#### FinnGen

Epilepsy was determined with ICD-10 G40, ICD-9 345, ICD-8 345 and Social Insurance Institution of Finland (KELA) code 111. Exclusion criteria were ICD-9 3452/3453 and ICD-8 34520. GGE was determined with ICD-10 G40.3, ICD-9 345[0-3] and ICD-8 34519. Exclusion criteria were ICD-8 34511. Focal epilepsy was determined with ICD-10 G40.0, G40.1, G40.2, ICD-9 345[45] and ICD-8 3453.

#### DeCode genetics

Epilepsy was determined with ICD-10 G40 and ICD-9 345 excluding 3452/3453. GGE with ICD-10 G40.3/G40.4/G40.6/G40.7 or ICD-9 3450/3451/3456, and focal epilepsy with ICD-10 G40.0/G40.1/G40.2 or ICD-9 3454/3455.

#### Biobank Japan

Cases were classified into “Broad_Epilepsy”, being any form of epilepsy; “Idiopathic_Epilepsy”, being epilepsy with onset under 40 years and no known cause; or “Idiopathic_Focal_Epilepsy” and “Idiopathic_Generalized_Epilepsy”, where focal and generalized syndromes could be ascertained.

Control data were population matched samples with no history of epilepsy. GWAS fixed-effects meta-analyses were conducted using METAL^67^. To account for case-control imbalance the effective sample size for each cohort was calculated as N_eff_ = 4/(1/N_cases_ + 1/N_controls_)). GWAS Manhattan plots were generated using the qqman R package^75^. Genome-wide significant loci were mapped onto genes using the FUMA web platform^18^.

We performed three meta-analyses. As a primary analysis, we meta-analysed all non-biobank samples, then we meta-analysed only biobank/deCODE genetics samples and finally performed a combined meta-analysis of biobank/deCODE genetics and non-biobank samples.

### Pleiotropy analysis

ASSET^76^ is a meta-analysis-based pleiotropy detection approach that identifies common or shared genetic effects between two or more related, but distinct traits. We used ASSET with a genome-wide significance level of α=5×10^−8^. We applied ASSET to the subset of European samples, comprising 6952 (3244+3708) GGE cases and 14,939 (5344+9095) focal epilepsy cases from the Epi25 and our Consortium as well as 42,434 partially overlapping controls from both consortia. Note that ASSET accounts for sample overlap in the analysis. Effect sizes, standard errors and the effective sample sizes estimated were from the main meta-analysis.

### HLA association

Given the prior association of the HLA with autoimmune epilepsy^77,78^, we included a specific analysis of the HLA. HLA types and amino acid residues were imputed using CookHLA software,^26^ with the 1000 Genomes Phase 3 used as a reference panel.^79^ Samples were grouped by genetic ancestry for imputation.

Following imputation, association analysis was conducted using the HLA Analysis Toolkit (HATK).^80^ Three phenotypes were analysed: ‘all epilepsy’, ‘focal epilepsy’ and ‘GGE’. Samples from the ILAE and Epi25 datasets were analysed separately and the association results were meta-analysed across datasets using PLINK.^81^

### Functional annotation

We annotated all genome-wide significant SNPs and tagged SNPs within the loci. ANNOVAR was used to retrieve the location and function of each SNP,^82^ the CADD score was used as a measure of predicted deleteriousness^83^ and chromatin states were incorporated from the ENCODE and NIH Roadmap Epigenomics Mapping Consortium.^14,84^ We used FUMA to define the independently significant SNPs within loci; i.e., SNPs that were genome-wide significant but not in LD (R^2^<0.2 in Europeans) with the lead SNP in the locus.

### Gene mapping

To map genome-wide significant loci to specific genes, we used FUMA^18^ with the same parameters as published previously.^4^ We defined genome-wide significant loci as the region encompassing all SNPs with P<10^−4^ that were in LD (R^2^>0.2) with the lead SNP (i.e., the SNP with the strongest association within the region). We used a combination of positional mapping (within 250 kb from the locus), eQTL mapping (SNPs with FDR corrected eQTL P<0.05 in blood or brain tissue) and 3D Chromatin Interaction Mapping (FDR p <10^−6^ in brain tissue).

### Genome-wide gene based association study and gene-set analyses

We performed the genome-wide gene based association study (GWGAS) using default settings of MAGMA v1.08, as implemented in FUMA, which calculates an association P-value based on all the associations of all SNPs within each gene in the GWAS.^19^ Based on these GWGAS results, we performed competitive gene-set analyses with default MAGMA settings, using 15,483 default gene sets and GO-terms from MsigDB. In addition, we specifically assessed 18 curated gene-sets involving different synaptic functions.^29^

### Transcriptome wide association study

Transcriptome wide association studies (TWAS) were performed with FUSION v3, with default settings.^20^ We imputed gene expression based on our European-only GWAS (since the method relies on LD reference data) eQTL data from the PsychENCODE consortium, which includes dorsolateral prefrontal cortex tissue from 1,695 human subjects.^21^

### Summary-data-based Mendelian Randomization

Summary-data-based Mendelian Randomization (SMR) v1.03 is an additional method to assess the association between epilepsy and expression of specific genes.^22^ SMR tests whether the effect size of a SNP on epilepsy is mediated by expression of specific genes. We performed SMR analyses with default settings, using the MetaBrain expression data as reference; a new eQTL dataset including 2,970 human brain samples.^85^

### Sex-specific analyses

We performed a GWAS as described above for all epilepsy (13,889 female cases and 19,676 female controls; 12,259 male cases and 18,645 male controls) and GGE (3,946 female cases and 19,676 female controls; 2,603 male cases and 18,645 male controls) separately for subjects of either sex, after which we performed fixed-effects meta-analyses with METAL to merge the different cohorts. We performed meta-analyses between the male and female GWAS with GWAMA^86^ to assess heterogeneity of effect sizes between sexes and gender-differentiated associations^.33^

### Gene prioritization

We combined 10 methods to prioritize the most likely biological candidate gene within each genome-wide significant locus. For each gene in each locus, we assessed the following criteria:

- Missense: we assessed whether the SNPs tagged in the genome-wide significant locus contained an exonic missense variant in the gene, as annotated by ANNOVAR.
- TWAS: we assessed whether imputed gene expression was significantly associated with the epilepsy phenotype, based on the FUSION TWAS as described above, Bonferroni corrected for each mapped gene with expression information.
- SMR: we assessed whether the gene had a significant SMR association with the epilepsy phenotype, based on the SMR analyses as described above, Bonferroni corrected for each mapped gene with expression information.
- MAGMA: we assessed whether the gene was significantly associated with the epilepsy phenotype through a GWGAS analysis, Bonferroni corrected for each mapped gene.
- PoPS: we calculated the Polygenic Priority Score (PoPS)^87^; a novel method that combines GWAS summary statistics with biological pathways, gene expression, and protein-protein interaction data, to pinpoint the most likely causal genes. We scored the gene with the highest PoPS score within each locus.
- Brain expression: we calculated mean expression of all brain and non-brain tissues based on data from the Genotype-Tissue Expression (GTEx) project v8^88^ and assessed if the average brain tissue expression was higher than the average expression in non-brain tissues.
- brain-coX: we assessed whether genes were prioritized as co-expressed with established epilepsy genes in more than a third of brain tissue resources utilized, using the tool brain-coX (**Supplementary figure 17**).^89^
- Target of AED: we assessed whether the gene is a known target of an anti-epileptic drug, as detailed in the drug-gene interaction database (www.DGidb.com; accessed on 26-11-2021) and a list of drug targets from a recent publication (**Supplementary data 9**).^90^
- Knockout mouse: we assessed whether a knockout of the gene in a mouse model results in a nervous system (phenotype ID: MP:0003631) or a neurological/behaviour phenotype (MP:0005386) in the Mouse Genome Informatics database (http://www.informatics.jax.org; accessed on 26-11-2021).
- Monogenic epilepsy gene: we evaluated whether the gene is listed as a monogenic epilepsy gene, in a curated list maintained by the Epilepsy Research Centre at the University of Melbourne (**Supplementary data 9**).

Similar to previous studies,^4,91^ we scored all genes based on the number of criteria being met (range 0-10; all criteria had an equal weight). The gene with the highest score was chosen as the most likely implicated gene. We implicated both genes if they had an identical, highest score.

### Long distance expression regulation of BCL11A

Most eQTL databases, like PsychENCODE and MetaBrain, restrict eQTL analyses to 1 MB distance between genes and SNPs. To specifically assess the hypothesis of long-distance regulation of *BCL11A* by the lead SNPs in the 2p16.1 epilepsy locus, we manually interrogated the MetaBrain database^85^ without distance restraints. Next, we calculated the association between the 3 lead SNPs in the locus (rs11688767, rs77876353, rs13416557) with BCL11A expression.

### Heritability analyses

We calculated SNP-based heritability on the European-only GWAS using LDAK with default settings and pre-calculated LD weights from 2000 European (white British) reference samples under the BLD-LDAK SumHer model, as recommended for human traits.^92^ SNP based heritabilities were converted to liability scale heritability estimates, using the formula: h2L=h2o∗K2(1−K)2/p(1−p)∗Z2, where K is the disease prevalence, p is the proportion of cases in the sample, and Z is the standard normal density at the liability threshold. To decrease downward bias, we performed these calculations based on the effective sample sizes (see calculation above), after which p=0.5 can be assumed,^93^ with the same population prevalences as our previous study.^4^

The total amount of causally associated variants (i.e., variants with nonzero additive genetic effect) underlying epilepsy risk was calculated by a causal mixture model (MiXeR).^36^ MiXeR utilizes a likelihood-based framework to estimate the amount of causal SNPs underlying a trait, without the need to pinpoint which specific SNPs are involved. Furthermore, MiXeR allows for power calculations to assess the required sample size to explain a certain proportion SNP-based heritability by genome-wide significant SNPs.

### Enrichment analyses

We used MAGMA (as implemented in FUMA) to perform tissue and cell-type enrichment. First, we assessed whether our GGE GWAS was enriched for specific tissues from the GTEx database. Similarly, we assessed enrichment of genes expressed in the brain at 11 general developmental stages, using data from the BrainSpan consortium. Next, we assessed whether GGE was associated with specific cell types, by cross-referencing two single-cell RNA sequencing databases of human developmental and adult brain samples. The PsychENCODE database contains RNA sequencing data from 4,249 human brain cells from developmental stages and 27,412 human adult brain cells.^94^ The Zhong dataset (GSE104276) contains RNA sequencing data from 2,309 human brain cells at different stages in development.^95^ We performed FDR correction across datasets to assess which cell types were significantly associated with GGE. As sensitivity analysis, we performed stratified LDSC with default settings using the cell-specific gene expression weights from the PsychENCODE consortium to compare GABAergic with glutamatergic neuron enrichment.^96^

### Genetic overlap with other diseases

Using the FUMA web application, we searched the GWAS Catalog for previously reported associations with P< 5×10^−8^ for SNPs at all 26 genome-wide significant loci.

Genetic correlations between all, focal epilepsy and GGE and other traits were computed with LDSC, using default settings. Traits highlighted by the GWAS catalog analysis and/or those with established epilepsy comorbidity were prioritized and pursued provided recent summary statistics were available for public download (**Supplementary table 11**).

We used a recently described bivariate causal mixture model to quantify polygenic overlap between GGE with intelligence and autism spectrum disorder (ASD). Publicly available summary statistics from intelligence (n=269867) and ASD GWAS (n=46350) were downloaded,^97,98^ after which bivariate MiXeR was run with default settings.

### Drug-repurposing analyses

We utilized a recently developed method that uses the GWAS for a disease to predict the relative efficacy of drugs for the disease.^42^ We applied this method to the all epilepsy and GGE GWAS results, using (1) imputed gene expression data from the FUSION analyses, as described above, and (2) gene-based p-values from MAGMA (see above), with default settings. We predicted the relative efficacy of 1343 drugs in total (**Supplementary data 8**). We determined if our predictions correctly identify (area under receiver operating characteristic curve) and prioritize (median rank) known clinically-effective antiseizure drugs, as previously described.^42^ We determined the statistical significance of drug identification and prioritization results by comparing the results to those from a null distribution generated by performing 10^6^ random permutations of the scores assigned to drugs.

### Data availability

The GWAS summary statistics data that support the findings of this study (for both trans-ethnic and European-only analyses) are available at https://www.epigad.org/.

## Author contributions

### Data analysis

*Analytical design, imputation:*O.M.Adesoji, M.Bahlo, C.Campbell (lead analyst), G.L.Cavalleri, S.Chen (lead analyst), Y-C.A.Feng, B.P.C.Koeleman, R.Krause (data management), D.Lal, C.Leu, N.Mirza, M.Nothnagel, K.L.Oliver, R.Stevelink (lead analyst).

*Data generation and quality control and management:* L.Baum, J.P.Bradfield, R.J.Buono, G.L.Cavalleri., F.Cerrato, S.S.Cherny, C.Churchhouse, C.Cusick, Y-C.A.Feng, N.Gupta, H.Hakonarson, E.L.Heinzen, I.Helbig, D.P.Howrigan, D.Kasperaviciute, B.P.C.Koeleman, R.Krause., D.Lal, Z.Landoulsi, C.Leu, I.Lopes-Cendes., P.May, N.Mirza, B.M.Neale, P.-W.Ng, P.Nürnberg, Sl.Petrovski, T.Sander, D.Speed, R.Stevelink, Fe.Zara, W.Zhou.

*External data resources and analysis:* UK BioBank: C.Campbell, D.Lewis-Smith, R.H.Thomas. BioBank Japan: Y.Kamatani, M.Kanai, M.Kato, Y.Okada.

FinnGenn: M.J.Daly, H.O.Heyne, R.Kälviäinen, M.I.Kurki, A.Palotie.

deCODE genetics: S.Magnusson, E.Ólafsson, H.Stefansson, K.Stefansson, U.Unnsteinsdóttir.

*Analysis coordination:* G.L.Cavalleri (Co-Chair), B.P.C.Koeleman (Co-Chair)

### Writing committee

O.M.Adesoji, M.Bahlo, S.F.Berkovic, C.Campbell, G.L.Cavalleri, S.Chen, B.P.C.Koeleman, K.L.Oliver, R.Stevelink (wrote first draft).

### Strategy committee

L.Baum, S.F.Berkovic (Chair), R.J.Buono, G.L.Cavalleri, H.Hakonarson, E.L.Heinzen, M.R.Johnson, R.Kalviainen, B.P.C.Koeleman, R.Krause, P.Kwan, D.Lal, H.Lerche, Q.S.Li, I.Lopes-Cendes, D.H.Lowenstein, T.J.O’Brien, S.M.Sisodiya.

### Phenotyping committee

C.Depondt, D.J.Dlugos, W.S.Kunz, P.Kwan, D.H.Lowenstein (Chair), A.G.Marson, P.Striano.

### Governance committee

S.F.Berkovic, A.Compston, A-E.Lehesjoki, D.H.Lowenstein.

### Patient recruitment and phenotyping

Z.Afawi, E.Amadori, A.Anderson, J.Anderson, D.M.Andrade, G.Annesi, A.Avbersek, M.D.Baker, G.Balagura, S.Balestrini, C.Barba, K.Barboza, F.Bartolomei, T.Bast, T.Baumgartner, B.Baykan, N.Bebek, A.J.Becker, F.Becker, C.A.Bennett, B.Berghuis, S.F.Berkovic, A.Beydoun, C.Bianchini, F.Bisulli, I.Blatt, I.Borggraefe, C.Bosselmann, V.Braatz, K.Brockmann, R.J.Buono, R.M.Busch, H.Caglayan, E.Campbell, L.Canafoglia, C.Canavati, G.D.Cascino, B.Castellotti, C.B.Catarino, F.Chassoux, K.Chinthapalli, I-J.Chou, S-K.Chung, P.O.Clark, A.J.Cole, A.Coppola, M.Cosico, P.Cossette, J.J.Craig, L.K.Davis, G-J.deHaan, N.Delanty, C.Depondt, P.Derambure, O.Devinsky, L.Di Vito, D.J.Dlugos, V.Doccini, C.P.Doherty, H.El-Naggar, C.E.Elger, C.A.Ellis, A.Faucon, L.Ferguson, T.N.Ferraro, L.Ferri, M.Feucht, M.Fitzgerald, B.Fonferko-Shadrach, F.Fortunato, S.Franceschetti, J.A.French, E.Freri, M.Gagliardi, A.Gambardella, E.B.Geller, T.Giangregorio, L.Gjerstad, T.Glauser, E.Goldberg, A.Goldman, T.Granata, D.A.Greenberg, R.Guerrini, K.Hallmann, M.Hegde, I.Helbig, C.Hengsbach, S.Hirose, E.Hirsh, H.Hjalgrim, P-C.Hung, M.Iacomino, L.L.Imbach, Y.Inoue, A.Ishii, J.Jamnadas-Khoda, L.Jehi, M.R.Johnson, R.Kälviainen, M.Kanaan, A.-M.Kantanen, B.Kara, S.M.Kariuki, D. Kasteleijn-Nolst Trenite, J.Kegele, Y.Kesim, N.Khoueiry-Zgheib, C.King, H.E.Kirsch, K.M.Klein, G. Kluger, S.Knake, R.C.Knowlton, A.D.Korczyn, A.Koupparis, I.Kousiappa, M.Krenn, H.Krestel, I.Krey, W.S.Kunz, G.Kurlemann, Ru.Kuzniecky, P.Kwan, A.Labate, A.Lacey, S.Lauxmann, S.L.Leech, A-E.Lehesjoki, J.R.Lemke, H.Lerche, G.Lesca, B.Neubauer, N.Lewin, Q.S.Li, L.Licchetta, K-L.Lin, D.Lindhout, T.Linnankivi, I.Lopes-Cendes, D.H.Lowenstein, C.H.T.Lui, F.Madia, A.G.Marson, C.M.McGraw, D.Mei, R.Minardi, R.S.Moller, M.Montomoli, B.Mostacci, L.Muccioli, H.Muhle, K.Müller-Schlüter, I.M.Najm, W.Nasreddine, C.R.J.C.Newton, T.J.O’Brien, Ç.Özkara, S.S.Papacostas, E.Parrini, M.Pendziwiat, W.O.Pickrell, R.Pinsky, T.Pippucci, An.Poduri, F.Pondrelli, R.H.W.Powell, M.Privitera, A.Rademacher, R.Radtke, F.Ragona, S.Rau, M.I.Rees, B.M.Regan, P.S.Reif, S.Rhelms, A.Riva, F.Rosenow, P.Ryvlin, A.Saarela, L.G.Sadleir, J.W.Sander, Th.Sander, M.Scala, Th.Scattergood, S.C.Schachter, C.J.Schankin, I.E.Scheffer, B.Schmitz, S.Schoch, S.Schubert-Bast, A.Schulze-Bonhage, P.Scudieri, B.R.Sheidley, J.J.Shih, G.J.Sills, S.M.Sisodiya, M.C.Smith, P.E.Smith, A.C.M.Sonsma, M.R.Sperling, B.J.Steinhoff, U.Stephani, W.C.Stewart, C.Stipa, P.Striano, H.Stroink, A.Strzelczyk, R.Surges, T.Suzuki, K.M.Tan, G.A.Tanteles, E.Tauboll, L.L.Thio, O.Timonen, P.Tinuper, M.Todaro, P.Topaloglu, R.Tozzi, M-H.Tsai, B.Tumiene, D.Turkdogan, A.Utkus, P.Vaidiswaran, L.Valton, A.van Baalen, A.Vetro, E.P.G.Vining, F.Visscher, S.von Brauchitsch, R.von Wrede, R.G.Wagner, Y.G.Weber, S.Weckhuysen, J.Weisenberg, M.Weller, C.D.Whelan, P.Widdess-Walsh, M.Wolff, S.Wolking, D.Wu, K.Yamakawa, Z.Yapici, E.Yücesan, S.Zagaglia, F.Zahnert, F.Zimprich, G.Zsurka, Q.Zulfiqar Ali.

### Control cohorts

L.C.Brody, J.G.Eriksson, A.Franke, H.Hakonarson, Y.-L.Lau, J.L.Mills, A.M.Molloy, M.M.Nöthen, A.Palotie, F.Pangilinan, H.Stroink, W.Yang.

### Consortium coordination

K.L.Oliver.

## Author names and affiliations

**The International League Against Epilepsy Consortium on Complex Epilepsies\***

^*^three lead analysts listed first followed by all members in alphabetical order

Remi Stevelink^1^, Ciarán Campbell^2, 3^, Siwei Chen^4, 5^, Oluyomi M Adesoji^6^, Zaid Afawi^7^, Elisabetta Amadori^8, 9^, Alison Anderson^10, 11^, Joseph Anderson^12^, Danielle M Andrade^13^, Grazia Annesi^14^, Andreja Avbersek^15^, Melanie Bahlo^16-18^, Mark D Baker^19^, Ganna Balagura^8, 9^, Simona Balestrini^15, 20^, Carmen Barba^21^, Karen Barboza^22^, Fabrice Bartolomei^23^, Thomas Bast^24, 25^, Larry Baum^26, 27^, Tobias Baumgartner^28^, Betül Baykan^29, 30^, Nerses Bebek^29, 30^, Albert J Becker^31^, Felicitas Becker^32^, Caitlin A Bennett^33^, Bianca Berghuis^34^, Samuel F Berkovic^33^, Ahmad Beydoun^35^, Claudia Bianchini^21^, Francesca Bisulli^36, 37^, Ilan Blatt^7, 38^, Ingo Borggraefe^39, 40^, Christian Bosselmann^41^, Vera Braatz^15, 20^, Jonathan P Bradfield^42, 43^, Knut Brockmann^44^, Lawrence C Brody^45^, Russell J Buono^42, 46, 47^, Robyn M Busch^48-50^, Hande Caglayan^51^, Ellen Campbell^52^, Laura Canafoglia^53^, Christina Canavati^54^, Gregory D Cascino^55^, Barbara Castellotti^56^, Claudia B Catarino^15^, Gianpiero L Cavalleri^2, 3^, Felecia Cerrato^57^, Francine Chassoux^58^, Stacey S Cherny^26, 59^, Krishna Chinthapalli^15^, I-Jun Chou^60^, Seo-Kyung Chung^61, 62^, Claire Churchhouse^4, 5, 57^, Peggy O Clark^63^, Andrew J Cole^64^, Alastair Compston^65^, Antonietta Coppola^66^, Mahgenn Cosico^67, 68^, Patrick Cossette^69^, John J Craig^70^, Caroline Cusick^57^, Mark J Daly^4, 5, 57, 71^, Lea K Davis^72-75^, Gerrit-Jan de Haan^76^, Norman Delanty^2, 3, 77^, Chantal Depondt^78^, Philippe Derambure^79^, Orrin Devinsky^80^, Lidia Di Vito^36^, Dennis J Dlugos^67^, Viola Doccini^21^, Colin P Doherty^3, 81^, Hany El-Naggar^2, 3, 77^, Christian E Elger^28^, Colin A Ellis^82^, Johan G Eriksson^83^, Annika Faucon^84^, Yen-Chen A Feng^4, 5, 57, 85, 86^, Lisa Ferguson^49^, Thomas N Ferraro^46, 87^, Lorenzo Ferri^36, 37^, Martha Feucht^88^, Mark Fitzgerald^67, 68, 82^, Beata Fonferko-Shadrach^19^, Francesco Fortunato^89^, Silvana Franceschetti^90^, Andre Franke^91^, Jacqueline A French^92^, Elena Freri^93^, Monica Gagliardi^14^, Antonio Gambardella^89^, Eric B Geller^94^, Tania Giangregorio^36^, Leif Gjerstad^95^, Tracy Glauser^63^, Ethan Goldberg^67, 68^, Alicia Goldman^96^, Tiziana Granata^93^, David A Greenberg^97^, Renzo Guerrini^21^, Namrata Gupta^5^, Hakon Hakonarson^42, 98^, Kerstin Hallmann^28, 99^, Manu Hegde^100^, Erin L Heinzen^101, 102^, Ingo Helbig^67, 68, 82, 91, 103, 104^, Christian Hengsbach^41^, Henrike O Heyne^5, 71, 105, 106^, Shinichi Hirose^107^, Edouard Hirsch^108^, Helle Hjalgrim^109, 110^, Daniel P Howrigan^4, 5, 57^, Po-Cheng Hung^60^, Michele Iacomino^9^, Lukas L Imbach^111^, Yushi Inoue^112^, Atsushi Ishii^113^, Jennifer Jamnadas-Khoda^15, 114^, Lara Jehi^49, 50^, Michael R Johnson^115^, Reetta Kälviäinen^116, 117^, Yoichiro Kamatani^118^, Moien Kanaan^54^, Masahiro Kanai^119, 120^, Anne-Mari Kantanen^116^, Bülent Kara^121^, Symon M Kariuki^122-124^, Dalia Kasperavičiute^15^, Dorothee Kasteleijn-Nolst Trenite^1^, Mitsuhiro Kato^125^, Josua Kegele^41^, Yeşim Kesim^29^, Nathalie Khoueiry-Zgheib^126^, Chontelle King^127^, Heidi E Kirsch^100^, Karl M Klein^128-131^, Gerhard Kluger^132, 133^, Susanne Knake^128, 131^, Robert C Knowlton^100^, Bobby P C Koeleman^1^, Amos D Korczyn^7^, Andreas Koupparis^134^, Ioanna Kousiappa^134^, Roland Krause^135^, Martin Krenn^136^, Heinz Krestel^129, 131, 137, 138^, Ilona Krey^139^, Wolfram S Kunz^28, 140^, Mitja I Kurki^4, 5, 57, 71^, Gerhard Kurlemann^141^, Ruben Kuzniecky^142^, Patrick Kwan^10, 11, 143^, Angelo Labate^144^, Austin Lacey^3, 77, 145^, Dennis Lal^48, 49, 57^, Zied Landoulsi^135^, Yu-Lung Lau^146^, Stephen Lauxmann^41^, Stephanie L Leech^33^, Anna-Elina Lehesjoki^147^, Johannes R Lemke^139^, Holger Lerche^41^, Gaetan Lesca^148^, Costin Leu^15, 48, 57^, Naomi Lewin^67, 68^, David Lewis-Smith^67, 104, 149, 150^, Qingqin S Li^151^, Laura Licchetta^36^, Kuang-Lin Lin^60^, Dick Lindhout^1, 76^, Tarja Linnankivi^152-154^, Iscia Lopes-Cendes^155^, Daniel H Lowenstein^100^, Colin H T Lui^156^, Francesca Madia^9^, Sigurdur Magnusson^157^, Anthony G Marson^158^, Patrick May^135^, Christopher M McGraw^64^, Davide Mei^21^, James L Mills^159^, Raffaella Minardi^36^, Nasir Mirza^158^, Rikke S Møller^109, 110^, Anne M Molloy^160^, Martino Montomoli^21^, Barbara Mostacci^36^, Lorenzo Muccioli^37^, Hiltrud Muhle^103^, Karen Müller-Schlüter^161^, Imad M Najm^49, 50^, Wassim Nasreddine^35^, Benjamin M Neale^4, 5, 57^, Bernd Neubauer^162^, Charles RJC Newton^122-124^, Markus M Nöthen^163^, Michael Nothnagel^6, 164^, Peter Nürnberg^6^, Terence J O’Brien^10, 11^, Yukinori Okada^120, 165^, Elías Ólafsson^166^, Karen L Oliver^16, 17, 33^, Çiğdem Özkara^167^, Aarno Palotie^4, 5, 57, 71^, Faith Pangilinan^45^, Savvas S Papacostas^134^, Elena Parrini^21^, Manuela Pendziwiat^91, 103^, Slavé Petrovski^10, 168^, William O Pickrell^19, 169^, Rebecca Pinsky^170^, Tommaso Pippucci^171^, Annapurna Poduri^170^, Federica Pondrelli^37^, Rob H W Powell^169^, Michael Privitera^172^, Annika Rademacher^103^, Rodney Radtke^173^, Francesca Ragona^93^, Sarah Rau^41^, Mark I Rees^62, 174^, Brigid M Regan^33^, Philipp S Reif^128, 129, 131^, Sylvain Rhelms^175, 176^, Antonella Riva^8, 9^, Felix Rosenow^128, 129, 131^, Philippe Ryvlin^177^, Anni Saarela^116, 117^, Lynette G Sadleir^127^, Josemir W Sander^15, 20, 76^, Thomas Sander^6, 178^, Marcello Scala^8, 9^, Theresa Scattergood^179^, Steven C Schachter^180^, Christoph J Schankin^137, 181^, Ingrid E Scheffer^33, 182^, Bettina Schmitz^178^, Susanne Schoch^31^, Susanne Schubert-Bast^129, 131^, Andreas Schulze-Bonhage^183^, Paolo Scudieri^8, 9^, Beth R Sheidley^170^, Jerry J Shih^184^, Graeme J Sills^185^, Sanjay M Sisodiya^15, 20^, Michael C Smith^186^, Philip E Smith^187^, Anja C M Sonsma^1^, Doug Speed^188, 189^, Michael R Sperling^190^, Hreinn Stefansson^157^, Kári Stefansson^157^, Bernhard J Steinhoff^24, 25^, Ulrich Stephani^103^, William C Stewart^191, 192^, Carlotta Stipa^36^, Pasquale Striano^8, 9^, Hans Stroink^193^, Adam Strzelczyk^128, 129, 131^, Rainer Surges^28^, Toshimitsu Suzuki^194, 195^, K Meng Tan^10^, George A Tanteles^134^, Erik Taubøll^95^, Liu Lin Thio^196^, Rhys H Thomas^149, 150^, Oskari Timonen^117^, Paolo Tinuper^36, 37^, Marian Todaro^10, 11^, Pınar Topaloğlu^197^, Rossana Tozzi^198^, Meng-Han Tsai^199^, Birute Tumiene^200, 201^, Dilsad Turkdogan^202^, Unnur Unnsteinsdóttir^157^, Algirdas Utkus^201^, Priya Vaidiswaran^67, 68^, Luc Valton^203^, Andreas van Baalen^103^, Annalisa Vetro^21^, Eileen P G Vining^204^, Frank Visscher^205^, Sophie von Brauchitsch^129, 131^, Randi von Wrede^28^, Ryan G Wagner^206^, Yvonne G Weber^41, 207^, Sarah Weckhuysen^208-210^, Judith Weisenberg^196^, Michael Weller^211^, Peter Widdess-Walsh^2, 3, 77^, Markus Wolff^212^, Stefan Wolking^207^, David Wu^84^, Kazuhiro Yamakawa^194, 195^, Wanling Yang^146^, Zuhal Yapıcı^197^, Emrah Yücesan^213^, Sara Zagaglia^15, 20^, Felix Zahnert^128^, Federico Zara^8, 9^, Wei Zhou^4, 5, 57^, Fritz Zimprich^136^, Gábor Zsurka^28, 140^, Quratulain Zulfiqar Ali^13^

1. Department of Genetics, University Medical Center Utrecht, Utrecht 3584 CX, The Netherlands.

2. School of Pharmacy and Biomolecular Sciences, The Royal College of Surgeons in Ireland, Dublin, Ireland.

3. The FutureNeuro Research Centre, Dublin, Ireland.

4. Analytic and Translational Genetics Unit, Department of Medicine, Massachusetts General Hospital and Harvard Medical School, Boston, MA 02114, USA.

5. Program in Medical and Population Genetics, Broad Institute of MIT and Harvard, Cambridge, MA, USA.

6. Cologne Center for Genomics (CCG), University of Cologne, Faculty of Medicine and University Hospital Cologne, 50931 Cologne, Germany.

7. Tel-Aviv University Sackler Faculty of Medicine, Ramat Aviv 69978, Israel.

8. Department of Neurosciences, Rehabilitation, Ophthalmology, Genetics, Maternal and Child Health, University of Genova, Genova, Italy.

9. IRCCS Istituto Giannina Gaslini, Genova, Italy.

10. Department of Medicine, University of Melbourne, Royal Melbourne Hospital, Parkville 3050, Australia.

11. Department of Neuroscience, Central Clinical School, Alfred Health, Monash University, Melbourne 3004, Australia.

12. Neurology Department, Aneurin Bevan University Health Board, Newport, Wales, UK.

13. Adult Genetic Epilepsy Program, University of Toronto, Toronto, ON, Canada.

14. Institute for Biomedical Research and Innovation, National Research Council, Cosenza, Italy.

15. Department of Clinical and Experimental Epilepsy, UCL Queen Square Institute of Neurology, London WC1N 3BG, UK.

16. Population Health and Immunity Division, The Walter and Eliza Hall Institute of Medical Research, Parkville 3052, Australia.

17. Department of Biology, University of Melbourne, Parkville 3010, Australia.

18. School of Mathematics and Statistics, University of Melbourne, Parkville 3010, Australia.

19. Swansea University Medical School, Swansea University, Swansea, Wales, UK.

20. Chalfont Centre for Epilepsy, Chalfont-St-Peter, Buckinghamshire SL9 0RJ, UK.

21. Pediatric Neurology, Neurogenetics and Neurobiology Unit and Laboratories, Children’s Hospital A. Meyer, University of Florence, Italy.

22. University Health Network, University of Toronto, Toronto, ON, Canada.

23. APHM, Timone Hospital, Epileptology and Cerebral Rhythmology, Aix Marseille Univ, INSERM, INS, Inst Neurosci Syst, Marseille, France.

24. Epilepsy Center Kork, Kehl-Kork 77694, Germany.

25. Medical Faculty of the University of Freiburg, Freiburg 79085, Germany.

26. Department of Psychiatry, The University of Hong Kong, Hong Kong.

27. The State Key Laboratory of Brain and Cognitive Sciences, University of Hong Kong, Hong Kong, China.

28. Department of Epileptology, University of Bonn Medical Centre, Bonn 53127, Germany.

29. Department of Neurology, Istanbul Faculty of Medicine, Istanbul University, Istanbul, Turkey.

30. Department of Genetics, Aziz Sancar Institute of Experimental Medicine, Istanbul University, Istanbul, Turkey.

31. Section for Translational Epilepsy Research, Department of Neuropathology, University of Bonn Medical Center, Bonn 53105, Germany.

32. Department of Neurology, University of Ulm, Ulm 89081, Germany.

33. Epilepsy Research Centre, University of Melbourne, Austin Health, Heidelberg 3084, Australia.

34. Stichting Epilepsie Instellingen Nederland (SEIN), Zwolle 8025 BV, The Netherlands.

35. Department of Neurology, American University of Beirut Medical Center, Beirut, Lebanon.

36. IRCCS Istituto delle Scienze Neurologiche di Bologna, Bologna, Italy.

37. Department of Biomedical and Neuromotor Sciences, University of Bologna, Bologna, Italy.

38. Department of Neurology, Sheba Medical Center, Ramat Gan, Israel.

39. Department of Pediatric Neurology, Dr von Hauner Children’s Hospital, Ludwig Maximilians University, Munchen, Germany.

40. Epilepsy Center Munich, Munich, Germany.

41. Department of Neurology and Epileptology, Hertie Institute for Clinical Brain Research, University of Tübingen, Tübingen 72076, Germany.

42. Center for Applied Genomics, The Children’s Hospital of Philadelphia, Philadelphia, PA 19104, USA.

43. Quantinuum Research LLC, Wayne, PA 19087, USA.

44. Children’s Hospital, Dept. of Pediatric Neurology, University Medical Center Göttingen, Göttingen, Germany.

45. National Human Genome Research Institute, National Institutes of Health, Bethesda, MD 20892, USA.

46. Department of Biomedical Sciences, Cooper Medical School of Rowan University Camden, NJ 08103, USA.

47. Department of Neurology, Thomas Jefferson University Hospital, Philadelphia, PA 19107, USA.

48. Genomic Medicine Institute, Lerner Research Institute, Cleveland Clinic, Cleveland, OH 44195, USA.

49. Cleveland Clinic Epilepsy Center, Neurological Institute, Cleveland Clinic, Cleveland, OH 44195, USA.

50. Department of Neurology, Neurological Institute, Cleveland Clinic, Cleveland, OH 44195, USA.

51. Department of Molecular Biology and Genetics, Bogaziçi University, Istanbul, Turkey.

52. Belfast Health and Social Care Trust, Belfast BT9 7AB, UK.

53. Integrated Diagnostics for Epilepsy, Fondazione IRCCS Istituto Neurologico C. Besta, Milan, Italy.

54. Hereditary Research Lab, Bethlehem University, Bethlehem, Palestine.

55. Division of Epilepsy, Department of Neurology, Mayo Clinic, Rochester, MN 55902, USA.

56. Unit of Genetics of Neurodegenerative and Metabolic Diseases, Fondazione IRCCS Istituto Neurologico Carlo Besta, Milan, Italy.

57. Stanley Center for Psychiatric Research, Broad Institute of Harvard and M.I.T., Cambridge, MA 02142, USA.

58. Hôpital Lariboisière, Dept of Neurosurgery-Paris-Cité University, Paris, France.

59. Department of Epidemiology and Preventive Medicine, School of Public Health, Sackler Faculty of Medicine, Tel Aviv University, Tel Aviv 6997801, Israel.

60. Department of Pediatric Neurology, Chang Gung Memorial Hospital, Linkou Branch, and College of Medicine, Chang Gung University, Taoyuan, Taiwan.

61. Kids Neuroscience Centre, Kids Research, Children Hospital at Westmead, Sydney, New South Wales, Australia.

62. Neurology Research Group, Swansea University Medical School, Faculty of Medicine, Health & Life Science, Swansea University, SA2 8PP, UK.

63. Cincinnati Children’s Hospital Medical Center, Cincinnati, Ohio, USA.

64. Neurology, Massachusetts General Hospital, Boston, MA, USA.

65. Department of Clinical Neurosciences, Cambridge Biomedical Campus, Cambridge CB2 0SL, UK.

66. Department of Neuroscience, Reproductive and Odontostomatological Sciences, University Federico II, Naples 80131, Italy.

67. Division of Neurology, Children’s Hospital of Philadelphia, Philadelphia, 3401 Civic Center Blvd,Philadelphia, PA 19104, USA.

68. The Epilepsy NeuroGenetics Initiative (ENGIN), Children’s Hospital of Philadelphia, Philadelphia, 3401 Civic Center Blvd, Philadelphia, PA 19104, USA.

69. Department of Neurosciences, Université de Montréal, Montréal, CA 26758, Canada.

70. Department of Neurology, Royal Victoria Hospital, Belfast Health and Social Care Trust, Grosvenor Road, Belfast BT12 6BA, UK.

71. Institute for Molecular Medicine Finland (FIMM), University of Helsinki, Helsinki 0014, Finland.

72. Division of Genetic Medicine, Department of Medicine, Vanderbilt University Medical Center, Nashville, TN, USA.

73. Department of Psychiatry and Behavioral Sciences, Vanderbilt University Medical Center, Nashville, TN, USA.

74. Department of Biomedical Informatics, Vanderbilt University Medical Center, Nashville, TN, USA.

75. Vanderbilt Genetics Institute, Vanderbilt University Medical Center, Nashville, TN, USA.

76. Stichting Epilepsie Instellingen Nederland (SEIN), Heemstede 2103 SW, The Netherlands.

77. Department of Neurology, Beaumont Hospital, Dublin D09 FT51, Ireland.

78. Department of Neurology, Hôpital Erasme, Université Libre de Bruxelles, Bruxelles 1070, Belgium.

79. Department of Clinical Neurophysiology, Lille University Medical Center, EA 1046, University of Lille.

80. Department of Neurology, New York University/Langone Health, New York NY, USA.

81. Neurology Department, St. James’s Hospital, Dublin D03 VX82, Ireland.

82. Department of Neurology, University of Pennsylvania, Perelman School of Medicine, Philadelphia, PA, 19104 USA.

83. Department of General Practice and Primary Health Care, University of Helsinki and Helsinki University Hospital, Helsinki 0014, Finland.

84. Human Genetics Training Program, Vanderbilt University, Nashville, TN, USA.

85. Psychiatric & Neurodevelopmental Genetics Unit, Department of Psychiatry, Massachusetts General Hospital and Harvard Medical School, Boston, MA 02114, USA.

86. Division of Biostatistics, Institute of Epidemiology and Preventive Medicine, College of Public Health, National Taiwan University, Taipei 100, Taiwan.

87. Department of Pharmacology and Psychiatry, University of Pennsylvania Perlman School of Medicine, Philadelphia, PA 19104, USA.

88. Department of Pediatrics and Neonatology, Medical University of Vienna, Vienna 1090, Austria.

89. Institute of Neurology, Department of Medical and Surgical Sciences, University “Magna Graecia”, Catanzaro, Italy.

90. Neurophysiology, Fondazione IRCCS Istituto Neurologico Carlo Besta, Milan, Italy.

91. Institute of Clinical Molecular Biology, Christian-Albrechts-University of Kiel, University Hospital Schleswig Holstein, Kiel 24105, Germany..

92. Department of Neurology, NYU School of Medicine, New York City, NY 10003, USA.

93. Department of Pediatric Neuroscience, Fondazione IRCCS Istituto Neurologico Carlo Besta, Milan, Italy.

94. Institute of Neurology and Neurosurgery at St. Barnabas, Livingston, NJ 07039, USA.

95. Department of Neurology, Division of Clinical Neuroscience, Rikshospitalet Medical Centre, University of Oslo, Oslo, Norway.

96. Department of Neurology, Baylor College of Medicine.

97. Department of Pediatrics, Nationwide Children’s Hospital, Columbia, Ohio, USA.

98. Division of Human Genetics, Department of Pediatrics, The Perelman School of Medicine, University of Pennsylvania, Philadelphia, PA 19104, USA.

99. Life and Brain Center, University of Bonn Medical Center, Bonn 53127, Germany.

100. Department of Neurology, University of California, San Francisco, CA 94143, USA.

101. Division of Pharmacotherapy and Experimental Therapeutics, Eshelman School of Pharmacy, University of North Carolina at Chapel Hill, Chapel Hill, NC, 27599, USA.

102. Department of Genetics, School of Medicine, University of North Carolina at Chapel Hill, Chapel Hill, NC, 27599, USA.

103. Department of Neuropediatrics, University Medical Center Schleswig-Holstein, Christian-Albrechts-University, 24105 Kiel, Germany.

104. Department of Biomedical and Health Informatics (DBHi), Children’s Hospital of Philadelphia, Philadelphia, PA, 19104 USA.

105. Hasso Plattner Institute, Digital Health Center, University of Potsdam, Germany.

106. Hasso Plattner Institute, Mount Sinai School of Medicine, NY, US.

107. General Medical Research Center, School of Medicine, Fukuoka University, Japan.

108. Department of Neurology, University Hospital of Strasbourg, Strasbourg, France.

109. Danish Epilepsy Centre, Dianalund 4293, Denmark.

110. Institute of Regional Health Services Research, University of Southern Denmark, Odense 5000, Denmark.

111. Swiss Epilepsy Center, Klinik Lengg, Zurich, Switzerland.

112. National Epilepsy Center, Shizuoka Institute of Epilepsy and Neurological Disorder, Shizuoka, Japan.

113. Department of Pediatrics, Fukuoka Sanno Hospital, Japan.

114. Department of Psychiatry and Applied Psychology, Institute of Mental Health University of Nottingham, Nottingham NG7 2TU, UK.

115. Division of Brain Sciences, Imperial College London, London SW7 2AZ, UK.

116. Kuopio Epilepsy Center, Neurocenter, Kuopio University Hospital, Kuopio 70210, Finland.

117. Institute of Clinical Medicine, University of Eastern Finland, Kuopio 70210, Finland.

118. Department of Computational Biology and Medical Sciences, Graduate School of Frontier Sciences, the University of Tokyo, Tokyo, Japan.

119. The Broad Institute of M.I.T. and Harvard, Cambridge, MA 02142, USA.

120. Department of Statistical Genetics, Osaka University Graduate School of Medicine, Suita, Japan.

121. Department of Child Neurology, Medical School, Kocaeli University, Kocaeli, Turkey.

122. Neuroscience Unit, KEMRI-Wellcome Trust Research Programme, Kilifi, Kenya.

123. Department of Public Health, Pwani University, Kilifi, Kenya.

124. Department of Psychiatry, University of Oxford, Oxford, UK.

125. Department of Pediatrics, Showa University School of Medicine, Epilepsy Medical Center, Showa University Hospital, 1-5-8 Hatanodai, Shinagawa-ku, Tokyo 142-8555, Japan.

126. Department of Pharmacology and Toxicology, American University of Beirut Faculty of Medicine, Beirut, Lebanon.

127. Department of Paediatrics and Child Health, University of Otago, Wellington, New Zealand.

128. Epilepsy Center Hessen-Marburg, Department of Neurology, Philipps University Marburg, Marburg, Germany.

129. Epilepsy Center Frankfurt Rhine-Main, Center of Neurology and Neurosurgery, Goethe University Frankfurt, Frankfurt, Germany.

130. Departments of Clinical Neurosciences, Medical Genetics and Community Health Sciences, Hotchkiss Brain Institute & Alberta Children’s Hospital Research Institute, Cumming School of Medicine, University of Calgary, Calgary, Alberta, Canada.

131. LOEWE Center for Personalized Translational Epilepsy Research (CePTER), Goethe University Frankfurt, Germany.

132. Neuropediatric Clinic and Clinic for Neurorehabilitation, Epilepsy Center for Children and Adolescents, Vogtareuth, Germany.

133. Research Institute Rehabilitation / Transition, / Palliation, PMU Salzburg, Austria.

134. Cyprus Institute of Neurology and Genetics, Nicosia, Cyprus.

135. Luxembourg Centre for Systems Biomedicine, University of Luxembourg, Esch-sur-Alzette L-4362, Luxembourg.

136. Department of Neurology, Medical University of Vienna, Vienna 1090, Austria.

137. Department of Neurology, Inselspital, Bern University Hospital, University of Bern, Bern 3010, Switzerland.

138. Yale School of Medicine, New Haven, CT 06510, USA.

139. Institute of Human Genetics, University of Leipzig Medical Center, Leipzig, Germany.

140. Institute of Experimental Epileptology and Cognition Research, Medical Faculty, University of Bonn, Bonn, Germany.

141. Bonifatius Hospital Lingen, Neuropediatrics Wilhelmstrasse 13, 49808 Lingen, Germany.

142. Department of Neurology, Hofstra-Northwell Medical School, New York, NY, USA.

143. Department of Medicine and Therapeutics, Chinese University of Hong Kong, Hong Kong, China.

144. Department of Biomedical and Dental Sciences, Morphological and Functional Images (BIOMORF), University of Messina, Messina, Italy.

145. The School of Pharmacy and Biomolecular Sciences, RCSI Dublin.

146. Department of Paediatrics and Adolescent Medicine, The University of Hong Kong, Hong Kong.

147. Folkhälsan Research Center and Medical Faculty, University of Helsinki, Helsinki 00290, Finland.

148. Department of Medical Genetics, Hospices Civils de Lyon and University of Lyon, Lyon, France.

149. Translational and Clinical Research Institute, Newcastle University, Newcastle Upon Tyne, UK.

150. Department of Clinical Neurosciences, Newcastle Upon Tyne Hospitals NHS Foundation Trust, Newcastle Upon Tyne, UK.

151. Neuroscience Department, Janssen Research & Development, LLC, 1125 Trenton-Harbourton Road, Titusville, NJ, 08560, USA.

152. Child Neurology, New Children’s Hospital, Helsinki, Finland.

153. Pediatric Research Center, University of Helsinki, Helsinki, Finland.

154. Helsinki University Hospital, Helsinki, Finland.

155. Department of Translational Medicine, School of Medical Sciences, University of Campinas (UNICAMP), and the Brazilian Institute of Neuroscience and Neurotecnology; Campinas, SP, Brazil.

156. Department of Medicine, Tseung Kwan O Hospital, Hong Kong.

157. deCODE genetics Sturlugata 8, IS-102, Reykjavík Iceland.

158. Department of Pharmacology and Therapeutics, University of Liverpool, Liverpool L69 3GL, UK.

159. Division of Intramural Population Health Research, Eunice Kennedy Shriver National Institute of Child Health and Human Development, National Institutes of Health, Bethesda, MD 20892, USA.

160. School of Medicine, Trinity College Dublin, Dublin 2, Ireland.

161. Epilepsy Center for Children, University Hospital Ruppin-Brandenburg, Brandenburg Medical School, 16816 Neuruppin, Germany.

162. Pediatric Neurology, University of Giessen, Germany.

163. Institute of Human Genetics, University of Bonn Medical Center, Bonn 53127, Germany.

164. University Hospital Cologne, Cologne, Germany.

165. Laboratory for Systems Genetics, RIKEN Center for Integrative Medical Sciences, Yokohama, Japan.

166. Department of Neurology, Landspitalinn University Hospital, Reykjavik, Iceland.

167. Istanbul University-Cerrahpaşa, Cerrahpaşa Medical Faculty, Department of Neurology, Istanbul, Turkey.

168. Centre for Genomics Research, Discovery Sciences, BioPharmaceuticals R&D, AstraZeneca, Cambridge CB2 0AA, UK.

169. Department of Neurology, Morriston Hospital, Swansea Bay University Bay Health Board, Swansea, Wales, UK.

170. Epilepsy Genetics Program, Division of Epilepsy and Clinical Neurophysiology, Department of Neurology, Boston Children’s Hospital, Boston, MA, USA.

171. IRCCS Azienda Ospedaliero-Universitaria di Bologna, Medical Genetics Unit, Bologna, Italy.

172. Department of Neurology, Gardner Neuroscience Institute, University of Cincinnati Medical Center, Cincinnati, OH 45220, USA.

173. Department of Neurology, Duke University School of Medicine, Durham, NC 27710, USA.

174. Faculty of Medicine & Health, University of Sydney, Sydney, New South Wales, Australia.

175. Department of Functional Neurology and Epileptology, Hospices Civils de Lyon and University of Lyon, France.

176. Lyon’s Neuroscience Research Center, INSERM U1028 / CNRS UMR 5292, Lyon, France.

177. Department of Clinical Neurosciences, Centre Hospitalo-Universitaire Vaudois, Lausanne, Switzerland.

178. Department of Neurology, Charité Universitaetsmedizin Berlin, Campus Virchow-Clinic, Berlin 13353, Germany.

179. Department of Endocrinology, Hospital of The University of Pennsylvania, Philadelphia, PA 19104, USA.

180. Departments of Neurology, Beth Israel Deaconess Medical Center, Massachusetts General Hospital, and Harvard Medical School, Boston, MA 02215, USA.

181. Department of Neurology, Ludwig Maximilians University, Munchen, Germany.

182. Department of Neurology, Royal Children’s Hospital, Parkville 3052, Australia.

183. Department of Epileptology, University Hospital Freiburg, Freiburg, Germany.

184. Department of Neurosciences, University of California, San Diego, La Jolla, CA 92037, USA.

185. School of Life Sciences, University of Glasgow, Glasgow G12 8QQ, UK.

186. Rush University Medical Center, Chicago, IL 60612, USA.

187. Department of Neurology, Alan Richens Epilepsy Unit, University Hospital of Wales, Cardiff CF14 4XW, UK.

188. UCL Genetics Institute, University College London, London WC1E 6BT, UK.

189. Aarhus Institute of Advanced Studies (AIAS), Aarhus University, 8000 Aarhus, Denmark.

190. Department of Neurology and Comprehensive Epilepsy Center, Thomas Jefferson University, Philadelphia, PA 19107, USA.

191. Department of Pediatrics, Ohio State University, Columbus, OH, USA.

192. The Research Institute, Nationwide Children’s Hospital, Columbus, OH, USA.

193. CWZ Hospital, 6532 SZ Nijmegen, The Netherlands.

194. Department of Neurodevelopmental Disorder Genetics, Institute of Brain Science, Nagoya City University Graduate School of Medical Science, Nagoya, Aichi, Japan.

195. Laboratory for Neurogenetics, RIKEN Center for Brain Science, Wako, Saitama, Japan.

196. Department of Neurology, Washington University School of Medicine, St. Louis, MO 63110, USA.

197. Department of Child Neurology, Istanbul Faculty of Medicine, Istanbul University, Istanbul, Turkey.

198. C. Mondino National Neurological Institute, Pavia 27100, Italy.

199. Department of Neurology, Kaohsiung Chang Gung Memorial Hospital, Kaohsiung, Taiwan.

200. Centre for Medical Genetics, Vilnius University Hospital Santaros Klinikos, Vilnius, Lithuania.

201. Institute of Biomedical Sciences, Faculty of Medicine, Vilnius University, Vilnius, Lithuania.

202. Department of Child Neurology, Medical School, Marmara University, Istanbul, Turkey.

203. Epilepsy Unit, Department of Neurology, Brain and Cognition Research Center - CerCo, CNRS, UMR5549, University Hospital and University of Toulouse, Paul Sabatier University, Toulouse, France.

204. Departments of Neurology and Pediatrics, The Johns Hopkins University School of Medicine, Baltimore, MD 21287, USA.

205. Department of Neurology, Admiraal De Ruyter Hospital, Goes 4462, The Netherlands.

206. MRC/Wits Rural Public Health & Health Transitions Research Unit (Agincourt), School of Public Health, Faculty of Health Sciences, University of the Witwatersrand, Johannesburg, South Africa.

207. Department of Neurology and Epileptology, University of Aachen, Aachen 52074, Germany.

208. Applied & Translational Neurogenomics Group, VIB Center for Molecular Neurology, VIB, Antwerp, Belgium.

209. Department of Neurology, Antwerp University Hospital, Edegem 2650, Belgium.

210. Translational Neurosciences, Faculty of Medicine and Health Science, University of Antwerp, Antwerp, Belgium.

211. Department of Neurology, University Hospital and University of Zurich, Zürich, Switzerland.

212. Department of Pediatric Neurology, Vivantes Hospital Neukölln, 12351 Berlin, Germany.

213. Bezmialem Vakif University, Institute of Life Sciences and Biotechnology, Istanbul, Turkey.

## Acknowledgements and funding

*Some of the data reported in this paper were collected as part of a project undertaken by the International League against Epilepsy (ILAE) and some of the authors are experts selected by the ILAE. Opinions expressed by the authors, however, do not necessarily represent the policy or position of the ILAE*.

This study received support from Science Foundation Ireland (SFI) (16/RC/3948), co-funded under the European Regional Development Fund, the Research Unit FOR-2715 of the German Research Foundation (MN: NO755/6-1, NO755/13-1), from Wellcome Trust (grant 084730), European Union’s Seventh Framework Programme (FP7/2007-2013) under grant agreement n°279062 (EpiPGX), The Muir Maxwell Trust and the Epilepsy Society, UK. Part of this work was undertaken at University College London Hospitals, which received a proportion of funding from the NIHR Biomedical

Research Centres funding scheme. RS and BPCK are supported by an ‘Vrienden WKZ’ fund 1616091 (MING). SFB and IES are supported by a National Health and Medical Research Council (NHMRC) of Australia Program Grant [1091593]. MB is supported by an NHMRC Investigator grant [APP1195236]. KLO is supported by an Australian Government Research Training Program Scholarship [APP533086] provided by the Australian Commonwealth Government and the University of Melbourne. DLS identified data from people predicted to have epilepsy form the UK Biobank while funded by a Wellcome Clinical PhD Fellowship on the 4ward North program [203914/Z/16/Z]. MRJ was supported by the UKRI MRC Award No: MR/S02638X/1 and by the NIHR Imperial Biomedical Research Centre (BRC). Fundação de Amparo à Pesquisa do Estado de São Paulo (FAPESP), Brazil, grant number (2013/07559-3). The funding bodies had no role in the study design, data collection, analysis, and interpretation, or in writing the manuscript.

We thank the Epi25 principal investigators, local staff from individual cohorts, and all of the patients with epilepsy who participated in research studies at local centers for making possible this global collaboration and resource to advance epilepsy genetics research. This work is part of the Centers for Common Disease Genomics (CCDG) program, funded by the National Human Genome Research Institute (NHGRI) The *Eunice Kennedy Shriver* National Institute of Child Health and Human Development, and the National Heart, Lung, and Blood Institute (NHLBI). CCDG-funded Epi25 research activities at the Broad Institute, including genomic data generation in the Broad Genomics Platform, were supported by NHGRI grant UM1 HG008895 (PIs: Eric Lander, Stacey Gabriel, Mark Daly, Sekar Kathiresan). The Genome Sequencing Program efforts were also supported by NHGRI grant 5U01HG009088-02. The content is solely the responsibility of the authors and does not necessarily represent the official views of the National Institutes of Health. We thank the Stanley Center for Psychiatric Research at the Broad Institute for supporting the genomic data generation efforts as well as aggregation of control samples and cohorts to contribute to the Epi25 GWAS analyses. In particular, the Genomic Psychiatry Cohort controls were genotyped on the GSA-MD v1.0 by the Broad Genomics Platform with funding from NIH grant U01MH105641 and the Stanley Center for Psychiatric Research, Broad Institute of MIT and Harvard. The FINRISK controls were part of the FINRISK studies supported by the THL (formerly KTL: National Public Health Institute) through budgetary funds from the government, with additional funding from institutions such as the Academy of Finland, the European Union, ministries and national and international foundations and societies to support specific research purposes. The collection of the Hong Kong Osteoporosis Study (HKOS) control samples was funded by the Bone Health Fund and Research Grants Council – Early Career Scheme (Project number: 27100416). Other control datasets included IBD NIDDK and samples from the Mass General Brigham (MGB) Biobank available from dbGaP under study accession number phs002018.v1.p1.

We want to acknowledge the participants and investigators of FinnGen study. The FinnGen project is funded by two grants from Business Finland (HUS 4685/31/2016 and UH 4386/31/2016) and the following industry partners: AbbVie Inc., AstraZeneca UK Ltd, Biogen MA Inc., Bristol Myers Squibb (and Celgene Corporation & Celgene International II Sàrl), Genentech Inc., Merck Sharp & Dohme Corp, Pfizer Inc., GlaxoSmithKline Intellectual Property Development Ltd., Sanofi US Services Inc., Maze Therapeutics Inc., Janssen Biotech Inc, Novartis AG, and Boehringer Ingelheim. Following biobanks are acknowledged for delivering biobank samples to FinnGen: Auria Biobank (www.auria.fi/biopankki), THL Biobank (www.thl.fi/biobank), Helsinki Biobank (www.helsinginbiopankki.fi), Biobank Borealis of Northern Finland (https://www.ppshp.fi/Tutkimus-ja-opetus/Biopankki/Pages/Biobank-Borealis-briefly-in-English.aspx), Finnish Clinical Biobank Tampere (www.tays.fi/en-US/Research_and_development/Finnish_Clinical_Biobank_Tampere), Biobank of Eastern Finland (www.ita-suomenbiopankki.fi/en), Central Finland Biobank (www.ksshp.fi/fi-FI/Potilaalle/Biopankki), Finnish Red Cross Blood Service Biobank (www.veripalvelu.fi/verenluovutus/biopankkitoiminta) and Terveystalo Biobank (www.terveystalo.com/fi/Yritystietoa/Terveystalo-Biopankki/Biopankki/). All Finnish Biobanks are members of BBMRI.fi infrastructure (www.bbmri.fi). Finnish Biobank Cooperative -FINBB (https://finbb.fi/) is the coordinator of BBMRI-ERIC operations in Finland. The Finnish biobank data can be accessed through the Fingenious® services (https://site.fingenious.fi/en/) managed by FINBB.

