## Supplemental material for "Genome-wide meta-analysis of over 29,000 people with epilepsy reveals 26 loci and subtype-specific genetic architecture"

### Supplementary Material

#### Supplementary Tables

*Supplementary table 1. Overview of novel cohorts, genotyping array and sample sizes. HK: Hong Kong; JPN: Japan.*

| Cohort | Genotyping platform | Ethnicity | Phenotype | Cases |
|---|---|---|---|---|
| **Epi25** | Illumina Infinium GSA | European | All epilepsy | 11544 |
| | | | GGE | 3153 |
| | | | Focal | 4523 |
| | | | Unclassified | 3868 |
| | | | Control | 13121 |
| | | European: Finnish | All epilepsy | 474 |
| | | | GGE | 91 |
| | | | Focal | 314 |
| | | | Unclassified | - |
| | | | Control | 986 |
| | | African | All epilepsy | 612 |
| | | | GGE | 276 |
| | | | Focal | 230 |
| | | | Unclassified | 106 |
| | | | Control | 3838 |
| | | Asian: HK | All epilepsy | 839 |
| | | | GGE | 55 |
| | | | Focal | 639 |
| | | | Unclassified | 145 |
| | | | Control | 594 |
| | | Asian: JPN | All epilepsy | 256 |
| | | | GGE | 63 |
| | | | Focal | 4 |
| | | | Unclassified | 189 |
| | | | Control | 211 |
| **Norwegian GenEpa** | Human610-Quadv1 | European | All epilepsy | 201 |
| | | | Focal | 201 |
| | | | Control | - |
| **Swiss GenEpa** | Illumina Human610-Quadv1 | European | All epilepsy | 231 |
| | | | Focal | 231 |
| | | | Control | 259 |

| | | | | |
|---|---|---|---|---|
| **Jansen Pharmaceuticals** | Illumina 1M | European | All epilepsy | 410 |
| | | | Focal | 410 |
| | | | Control | 3016 |
| **Austrian GenEpa** | Illumina Human CNV 370 duo | European | All epilepsy | 165 |
| | | | Focal | 165 |
| | | | Control | 337 |

*Supplementary table 2. Overview of number of cases and controls, stratified by phenotype and ethnicity.*

| **Phenotype** | **Sub-phenotype description** | **n** | **EUR** | **ASI** | **AFR** |
|---|---|---|---|---|---|
| GGE | Generalized Epilepsy, not otherwise specified, with spike and wave EEG | 3352 | 3024 | 44 | 284 |
| | Childhood Absence Epilepsy (CAE) | 1072 | 1049 | 6 | 17 |
| | Juvenile Absence Epilepsy (JAE) | 671 | 662 | 4 | 5 |
| | Juvenile Myoclonic Epilepsy (JME) | 1813 | 1732 | 61 | 20 |
| | GTCS only, with spike and wave EEG | 499 | 485 | 3 | 11 |
| | **Subtotal** | **7407** | **6952** | **118** | **337** |
| Focal | Focal Epilepsy, not otherwise specified | 3981 | 3688 | 140 | 153 |
| | Focal Epilepsy, documented lesion negative | 6367 | 5778 | 466 | 123 |
| | Focal Epilepsy, documented hippocampal sclerosis (HS) | 1375 | 1260 | 107 | 8 |
| | Focal Epilepsy, documented lesion other than HS | 4661 | 4213 | 416 | 32 |
| | **Subtotal** | **16384** | **14939** | **1129** | **316** |
| Unclassified | Epilepsy, not otherwise specified | 6153 | 5668 | 379 | 106 |
| | **Cases** | **29944** | **27559** | **1626** | **759** |
| | **Controls** | **52538** | **42436** | **3680** | **6422** |
| | **Total subjects** | **82482** | **69995** | **5306** | **7181** |

*Supplementary table 3. Summary of all genome-wide significant loci including genomic position and independent significant SNPs. We defined the locus position as the region encompassing all SNPs with P<10-4 that were in LD ($R^2$>0.2) with the lead SNP.*

| **Phenotype** | **Locus name** | **Locus position (hg19)** | **Lead SNP** | **Number of independent significant SNPs** | **Independent significant SNPs** |
|---|---|---|---|---|---|
| All epilepsy | 2p16.1 | chr2:57917222-58505679 | rs13032423 | 1 | rs13032423 |
| | 2q24.3 | chr2:166716305-167124221 | rs59237858 | 2 | rs59237858; rs1960242 |
| | 9q21.13 | chr9:76297313-76625089 | rs4744696 | 1 | rs4744696 |
| | 10q24.32 | chr10:103493226-103989812 | rs3740422 | 1 | rs3740422 |
| GGE | 1q43 | chr1:237846053-237908911 | rs876793 | 1 | rs876793 |
| | 2p16.1 | chr2:57917222-58756729 | rs11688767 | 3 | rs11688767; rs77876353; rs13416557 |
| | 2q12.1 | chr2:104056769-104481325 | rs62151809 | 1 | rs62151809 |
| | 2q24.3 | chr2:166818404-166994996 | rs11890028 | 1 | rs11890028 |
| | 2q32.2 | chr2:191504467-191710069 | rs6721964 | 1 | rs6721964 |
| | 3p22.3 | chr3:36218075-36345769 | rs9861238 | 1 | rs9861238 |

| | | | | | |
|---|---|---|---|---|---|
| | 3p21.31 | chr3:50184538-50421081 | rs739431 | 1 | rs739431 |
| | 4p15.1 | chr4:31107765-31204950 | rs1463849 | 1 | rs1463849 |
| | 5q22.3 | chr5:113837198-114440966 | rs4596374 | 1 | rs4596374 |
| | 5q31.2 | chr5:136459562-136684519 | rs2905552 | 1 | rs2905552 |
| | 6q22.33 | chr6:128302874-128333682 | rs13219424 | 1 | rs13219424 |
| | 7p14.1 | chr7:41334517-41411165 | rs37276 | 1 | rs37276 |
| | 9q21.32 | chr9:86320233-86694759 | rs2780103 | 1 | rs2780103 |
| | 10q24.32 | chr10:103493226-103989812 | rs11191156 | 1 | rs11191156 |
| | 12q13.13 | chr12:52319584-52348259 | rs114131287 | 2 | rs4762030; rs10431492 |
| | 16p13.3 | chr16:7285674-7442293 | rs62014006 | 1 | rs62014006 |
| | 17p13.1 | chr17:8036060-8219478 | rs2585398 | 1 | rs2585398 |
| | 17q21.32 | chr17:45938105-46554456 | rs16955463 | 1 | rs16955463 |
| | 19p13.3 | chr19:2102543-2136680 | rs75483641 | 1 | rs75483641 |
| | 21q21.1 | chr21:21655062-21719113 | rs1487946 | 1 | rs1487946 |
| | 21q22.1 | chr21:32036541-32203274 | rs7277479 | 1 | rs7277479 |
| | 22q13.32 | chr22:48615721-48639993 | rs469999 | 1 | rs469999 |
| CAE | 2p16.1 | chr2:57942325-58484172 | rs12185644 | 1 | rs12185644 |
| JME | 4p12 | chr4:46250605-46397617 | rs17537141 | 1 | rs17537141 |
| | 8q23.1 | chr8:109733213-109922163 | rs3019359 | 1 | rs3019359 |
| | 16p11.2 | chr16:30603521-31275374 | rs1046276 | 1 | rs1046276 |

*Supplementary table 4. Results from ASSET pleiotropy analyses for the 4 all epilepsy loci. The associated phenotype/s in ASSET reflect the phenotypes driving the ASSET signal, which could be both GGE individually. *Evidence for pleiotropy between GGE and Focal epilepsy.*

| SNP (Risk allele) | Chr. | Locus | P-value (GGE) | P-value (FE) | OR (ASSET) | P-value (ASSET) | Associated phenotype in ASSET |
|---|---|---|---|---|---|---|---|
| *rs60055328(C) | 2 | 2q24.3 | 1.04e-7 | 9.62e-7 | 1.07 | 2.8e-10 | GGE, FE |
| *rs4744696(G) | 9 | 9q21.13 | 3.07e-7 | 8.63e-5 | 0.93 | 4.4e-8 | GGE, FE |
| rs13032423(G) | 2 | 2p16.1 | 2.88e-17 | 2.93e-3 | 0.85 | 8.9e-17 | GGE only |
| rs3740422(G) | 10 | 10q24.32 | 1.02e-13 | 4.07e-3 | 1.15 | 2.48e-12 | GGE only |

*Supplementary table 5. Heritability enrichment of 26 functional categories, as assessed with LDAK heritability enrichment analyses. Statistical significance (in bold) is defined as P<0.05/26=0.0019.*

| Annotation | Share | SD | Expected | Enrichment | SD | Z-score | P-value |
|---|---|---|---|---|---|---|---|
| Coding_UCSC | 0.069948 | 0.027717 | 0.016079 | 4.350169 | 1.723782 | 1.943499 | 0.051956 |
| Conserved_LindbladToh | 0.132451 | 0.042109 | 0.028542 | 4.640618 | 1.475346 | 2.467637 | 0.013601 |
| CTCF_Hoffman | 0.017421 | 0.041158 | 0.023919 | 0.728312 | 1.720706 | 0.157893 | 0.874541 |
| DGF_ENCODE | 0.166208 | 0.095467 | 0.138091 | 1.203615 | 0.691336 | 0.294524 | 0.768358 |
| DHS_Trynka | 0.139003 | 0.100726 | 0.167956 | 0.827614 | 0.599718 | 0.287445 | 0.773772 |

| | | | | | | | |
|---|---|---|---|---|---|---|---|
| Enhancer_Andersson | -0.00046 | 0.018624 | 0.004432 | -0.104203 | 4.202414 | 0.262754 | 0.79274 |
| Enhancer_Hoffman | 0.065675 | 0.041653 | 0.042964 | 1.528598 | 0.969479 | 0.545239 | 0.585589 |
| FetalDHS_Trynka | 0.124737 | 0.077198 | 0.085617 | 1.456925 | 0.901664 | 0.506758 | 0.612325 |
| H3K27ac_Hnisz | 0.466327 | 0.030543 | 0.393028 | 1.186496 | 0.077713 | 2.399804 | 0.016404 |
| H3K27ac_PGC2 | 0.350543 | 0.056076 | 0.272914 | 1.284446 | 0.20547 | 1.384368 | 0.166246 |
| H3K4me1_Trynka | 0.578187 | 0.066915 | 0.428916 | 1.34802 | 0.156009 | 2.230769 | 0.025696 |
| H3K4me3_Trynka | 0.229663 | 0.052572 | 0.137162 | 1.674388 | 0.38328 | 1.759518 | 0.07849 |
| H3K9ac_Trynka | 0.253873 | 0.05353 | 0.129088 | 1.966671 | 0.414683 | 2.331108 | 0.019748 |
| Intron_UCSC | 0.45682 | 0.026361 | 0.394342 | 1.158437 | 0.066848 | 2.370108 | 0.017783 |
| PromoterFlanking_Hoffman | 0.021395 | 0.026356 | 0.008596 | 2.488946 | 3.066092 | 0.485617 | 0.627239 |
| Promoter_UCSC | 0.083972 | 0.035858 | 0.048118 | 1.74512 | 0.745213 | 0.999875 | 0.317371 |
| Repressed_Hoffman | 0.41399 | 0.06557 | 0.452761 | 0.914367 | 0.144823 | 0.591294 | 0.554323 |
| SuperEnhancer_Hnisz | 0.242302 | 0.019093 | 0.169819 | 1.42682 | 0.112434 | 3.796183 | **0.000147** |
| TFBS_ENCODE | 0.225196 | 0.07818 | 0.133056 | 1.692483 | 0.58757 | 1.178554 | 0.238576 |
| Transcr_Hoffman | 0.420967 | 0.057675 | 0.35335 | 1.191361 | 0.163225 | 1.172376 | 0.241046 |
| TSS_Hoffman | 0.046568 | 0.02996 | 0.018755 | 2.482921 | 1.5974 | 0.928334 | 0.353234 |
| UTR_3_UCSC | 0.013363 | 0.017055 | 0.011944 | 1.118787 | 1.427882 | 0.083191 | 0.9337 |
| UTR_5_UCSC | 0.02092 | 0.016217 | 0.005928 | 3.529243 | 2.735834 | 0.924487 | 0.355233 |
| WeakEnhancer_Hoffman | -0.03501 | 0.039502 | 0.021357 | -1.63943 | 1.849623 | 1.42701 | 0.153577 |
| Super_Enhancer_Vahedi | 0.029165 | 0.008124 | 0.021624 | 1.348773 | 0.375709 | 0.928306 | 0.353249 |
| Typical_Enhancer_Vahedi | 0.026998 | 0.011727 | 0.022194 | 1.216444 | 0.528371 | 0.409644 | 0.682067 |

*Supplementary table 6. Estimation of inflation factor and the LD-score regression intercept stratified by phenotype. λ: genomic inflation factor, $\lambda_{1000}$: genomic inflation factor corrected for an equivalent study of 1000 cases and 1000 controls.*

| Phenotype | λ | $\lambda_{1000}$ | Mean $\chi 2$ | LDSC intercept |
|---|---|---|---|---|
| All epilepsy | 1.25 | 1.01 | 1.27 | 1.10 |
| Focal epilepsy | 1.17 | 1.01 | 1.17 | 1.10 |
| GGE | 1.26 | 1.02 | 1.35 | 1.04 |

*Supplementary table 7. SNP-based heritabilities as calculated by LDAK, with the BLD-LADK model. Observed-scale heritability is calculated using effective-sample sizes, after which it was converted to liability-scale heritability using the same prevalence estimates as our previous GWAS.[1]*

| Phenotype | cases | controls | K: prevalence | Z | Observed-scale heritability | Liability scale heritability |
|---|---|---|---|---|---|---|
| All epilepsy | 27559 | 42436 | 0.005 | 0.0145 | 0.3733 | 0.177 (0.155 - 0.199) |
| Focal epilepsy | 14939 | 42436 | 0.003 | 0.0091 | 0.3733 | 0.160 (0.140 - 0.180) |
| GGE | 6952 | 42436 | 0.002 | 0.0063 | 0.9955 | 0.395 (0.343 - 0.446) |
| JME | 1728 | 37339 | 0.00035 | 0.0013 | 2.1135 | 0.635 (0.510 - 0.760) |
| JAE | 662 | 37339 | 0.00015 | 0.0006 | 3.3528 | 0.900 (0.633 - 1.166) |
| CAE | 1049 | 37339 | 0.00015 | 0.0006 | 3.0427 | 0.816 (0.638 - 0.995) |
| GTCSA | 485 | 37339 | 0.0002 | 0.0008 | 1.7824 | 0.496 (0.140 - 0.853) |
| Focal HS | 1260 | 37339 | 0.00075 | 0.0026 | 1.4020 | 0.472 (0.294 - 0.649) |
| Focal other lesion | 4213 | 37339 | 0.00135 | 0.0044 | 0.6778 | 0.251 (0.188 - 0.313) |
| Focal non-lesional | 5778 | 37339 | 0.0009 | 0.0031 | 0.2452 | 0.085 (0.046 -0.124) |

*Supplementary table 8. Top 20 drugs that are licensed for conditions other than epilepsy, but are predicted to be efficacious for GGE, and have published evidence of antiseizure efficacy from multiple published studies and in multiple animal models. We do not advise immediate use of these drugs for people with epilepsy, prior to any clinical trials. AUD: audiogenic; electro: maximal electroshock; Kin: kindling; PTZ: pentylenetetrazol. Drugs are listed in alphabetical order.*

| Drug | Current indication | Studies' PubMed IDs | Models |
|---|---|---|---|
| Aspirin | Pain; pyrexia; antiplatelet | 11883156, 14671677, 16844276, 22765917, 28060522 | Pilo, PTZ, Electro |
| Biperiden | Parkinson's disease | 738231, 2858579 | Electro, other |
| Captopril | Hypertension; chronic heart failure; diabetic nephropathy | 2824310, 22107891, 25573423 | PTZ, AUD, Other |
| Citalopram | Depression; panic disorder | 21531632, 21962757, 22429158, 22578701 | KA, PILO, PTZ |
| Dapsone | Leprosy; dermatitis herpetiformis | 1817960, 7970237, 23729301 | KA, Kin |
| Dextromethorphan | Pain; addiction; cough | 1456842, 2079649, 2574061, 2666123, 2676564, 2806362, 3044591, 3374269, 3380326, 3768695, 8058587, 8094234, 8405092, 8856734, 9179861, 9187330, 10080248, 11182165, 12479976, 12586225, 15084442, 15723099 | KA, PTZ, Electro, AUD, Kin, other |
| Diltiazem | Angina; hypertension | 2272645, 7681002, 8152336, 22661180 | KA, PTZ, Electro |
| Doxepin | Depression; pruritus | 1456842, 19443935 | PTZ, Electro, other |
| Fluoxetine | Depression; bulimia nervosa; obsessive-compulsive disorder | 7999524, 8149989, 8384110, 8538363, 8816259, 9696406, 15680343, 16531634, 17215106, 23530452, 25754610 | Electro, AUD, other |
| Isradipine | Hypertension | 8118482, 9595291 | Electro, AUD |
| Lovastatin | Hypercholesterolaemia | 21224519, 23253428, 23352156 | KA, AUD |
| Nicardipine | Angina; hypertension | 7681002, 8152336, 8872866, 10608279, 11742591 | KA, PTZ, Kin, other |
| Nifedipine | Angina; hypertension; Raynaud's phenomenon; premature labour | 1628595, 1698518, 1747472, 1865996, 1946038, 2085727, 2272645, 2713089, 2744396, 7681002, 7694769, 8054599, 8118482, 8152336, 8474621, 8707372, 12126870, 12536054, 16573711, 20113637, 22661180, 22801414 | KA, PTZ, Electro, AUD, Kin, other |
| Nimodipine | Subarachnoid haemorrhage | 1628595, 1698518, 2272645, 2310938, 2463174, 2662221, 3784769, 7681002, 8152336, 8156970, 8156971, 8707372, 9389584, 9570719, 9689485, 10683952, 12372903, 12536054, 12539272, 15123017, 17193898, 17344939, 19761108, 23761887, 25225705, 25445375 | KA, Pilo, PTZ, Electro, AUD, Kin, other |
| Orphenadrine | Parkinsonism | 2624511, 19815957 | PTZ, Electro |
| Pimozide | Schizophrenia | 2272645, 6141554, 7875556 | PTZ, Electro, AUD, other |
| Pioglitazone | Diabetes mellitus | 20599832, 22436324, 27527983 | PTZ, other |
| Quetiapine | Schizophrenia; mania; depression | 21168466, 26188240 | PTZ, AUD |
| Tamoxifen | Breast cancer; anovulatory infertility | 12139106, 24903749 | Electro, Kin |
| Thalidomide | Malignant disease; immunosuppression | 17449064, 21592729, 24735834 | PTZ, Kin |

*Supplementary table 9. Genetic correlations between our main GWAS and Biobank GWAS (including deCODE genetics). P-values are shown, with standard errors in brackets.*

| | Primary All epilepsy | Primary Focal | Primary GGE |
|---|---|---|---|
| **Biobank All epilepsy** | 0.74 (0.106) | 0.5525 (0.1781) | 0.7036 (0.0879) |
| **Biobank Focal** | 0.5835 (0.1596) | 0.7637 (0.2505) | 0.4331 (0.1275) |
| **Biobank Gen** | 0.6231 (0.1434) | 0.307 (0.2176) | 0.6521 (0.1373) |

*Supplementary table 10. Sample sizes of the included Biobanks and deCODE genetics.*

| Cohort | All epilepsy | Focal | GGE | Controls |
|---|---|---|---|---|
| **UK Biobank** | 7,006 | - | - | 179,763 |
| **Japan Biobank** | 612 | 145 | 283 | 176,694 |
| **DECODE genetics** | 3,762 | 405 | 1,342 | 335,389 |
| **FinnGen** | 10,354 | 5,922 | 1,160 | 332,143 |
| **Total** | 21,734 | 6,472 | 2,785 | 1,023,989 |

*Supplementary table 11. Phenotypes and associated publications assessed for genetic correlations with epilepsy using LDSC.*

| Broad trait | Trait | Publication | Notes |
|---|---|---|---|
| Psychiatric | Bipolar disorder | Mullins *et al* 2021[2] | |
| Psychiatric | ADHD | Demontis *et al* 2019[3] | |
| Psychiatric | ASD | Grove *et al* 2019[4] | |
| Psychiatric | Schizophrenia | Trubetskoy *et al* 2022[5] | |
| Psychiatric | Depression | Howard *et al* 2019[6] | exc. UKBB and 23andMe |
| Neurological | Febrile seizures | Skotte *et al* 2022[7] | |
| Neurological | Parkinson's disease | Nalls *et al* 2019[8] | exc. 23andMe |
| Neurological | Alzheimer's disease | Wightman *et al* 2021[9] | exc. 23andMe |
| Neurological | Stroke | Malik *et al* 2018[10] | |
| Neurological | Headache | Meng *et al* 2018[11] | |
| Neurological / Autoimmune | Multiple sclerosis | International Multiple Sclerosis Genetics Consortium 2019[12] | |
| Autoimmune | Type 1 diabetes | Chiou *et al* 2021[13] | |
| Autoimmune | Systemic lupus erythematosus | Morris *et al* 2016[14] | |
| Cognitive | Intelligence | Savage, Jansen *et al* 2018[15] | |
| Sleep | Insomnia | Jansen *et al* 2019[16] | exc. 23andMe |
| Smoking | Ever smoked | Karlsson Linnér *et al* 2019[17] | |
| Metabolic | Type 2 diabetes | Mahajan *et al* 2018[18] | |
| Metabolic | Coronary disease | van der Harst, Verweij *et al* 2018[19] | |

### Supplementary Figures

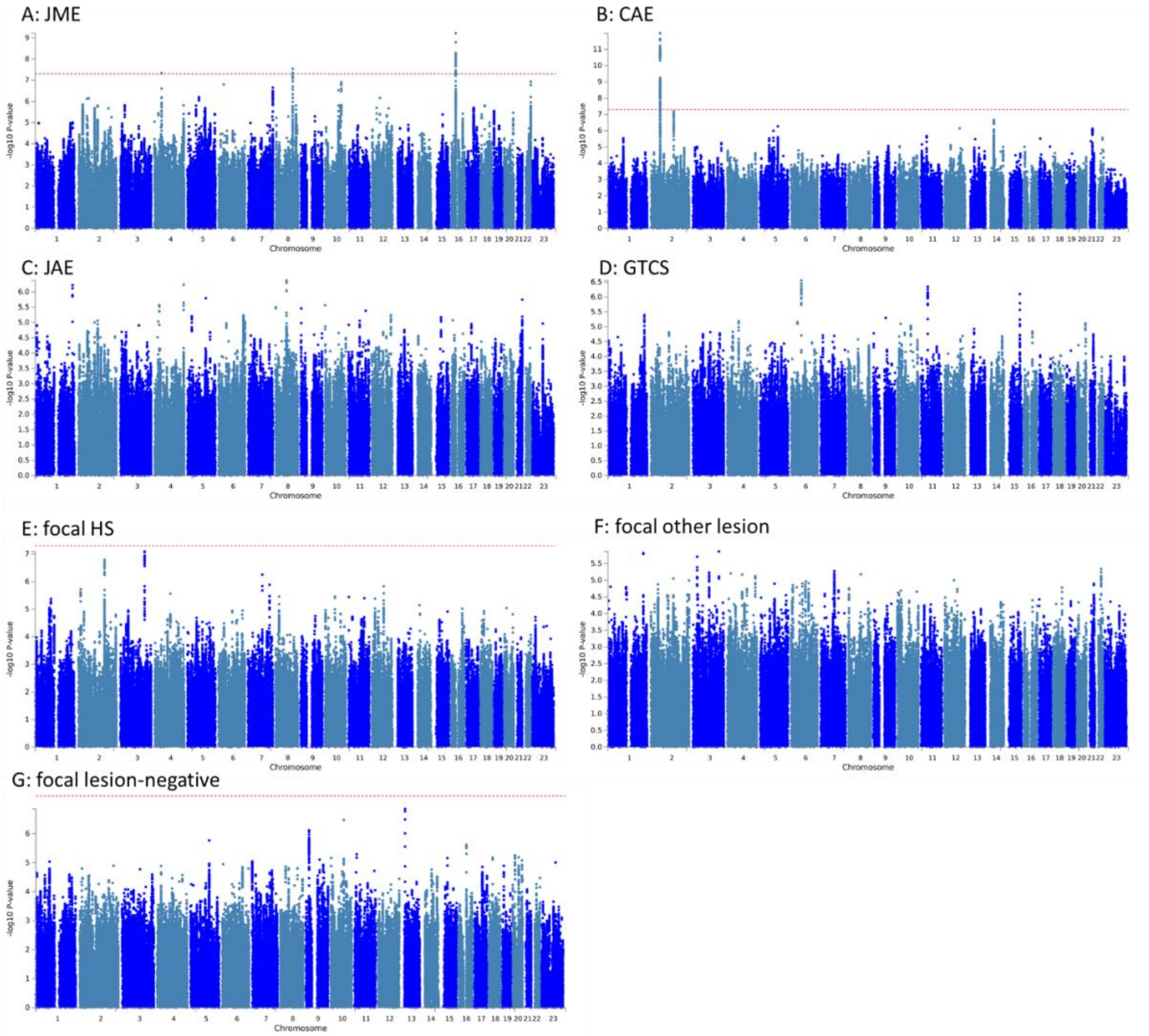


*Supplementary figure 1. Manhattan plots of epilepsy subphenotype GWAS. Chromosomal position is plotted on the X-axis and -log10 transformed P-values are plotted on the Y-axis. A. juvenile myoclonic epilepsy (JME); B. childhood absence epilepsy (CAE); C. juvenile absence epilepsy (JAE); D. generalized tonic-clonic seizures alone (GTCS); E. focal epilepsy due to hippocampal sclerosis (focal HS); F. focal epilepsy with other lesion; G. lesion negative focal epilepsy.*

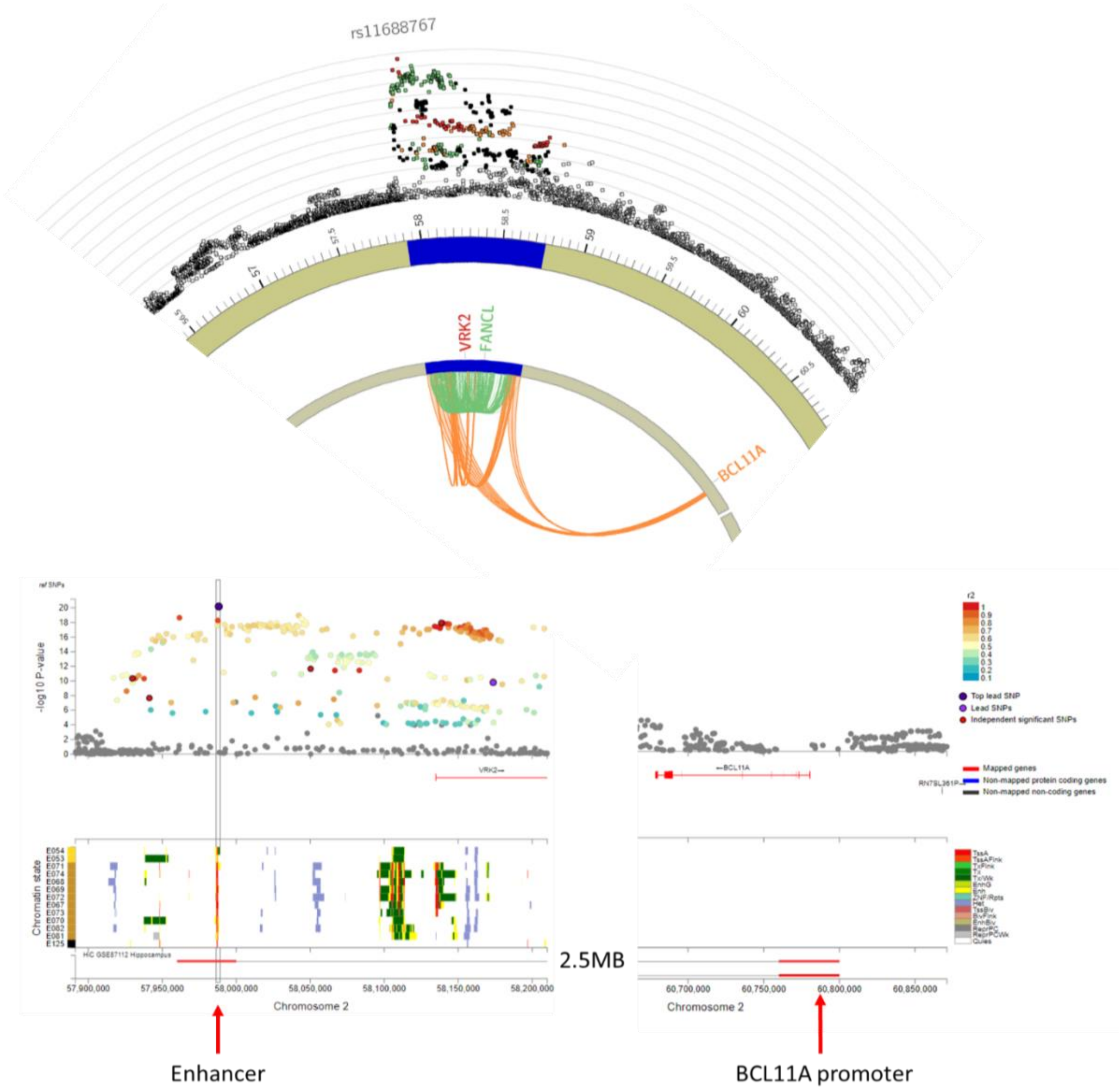


*Supplementary figure 2. 3D chromatin interactions link the 2p16.1 locus with the promoter region of BCL11A. The upper circus plot shows the 2p16.1 locus with GWAS P-values in the outer ring, with eQTL associations in green and HiC 3D chromatin interactions in orange. The locuszoom below shows GWAS P-values with chromatin states and Hi-C chromatin interactions below.*

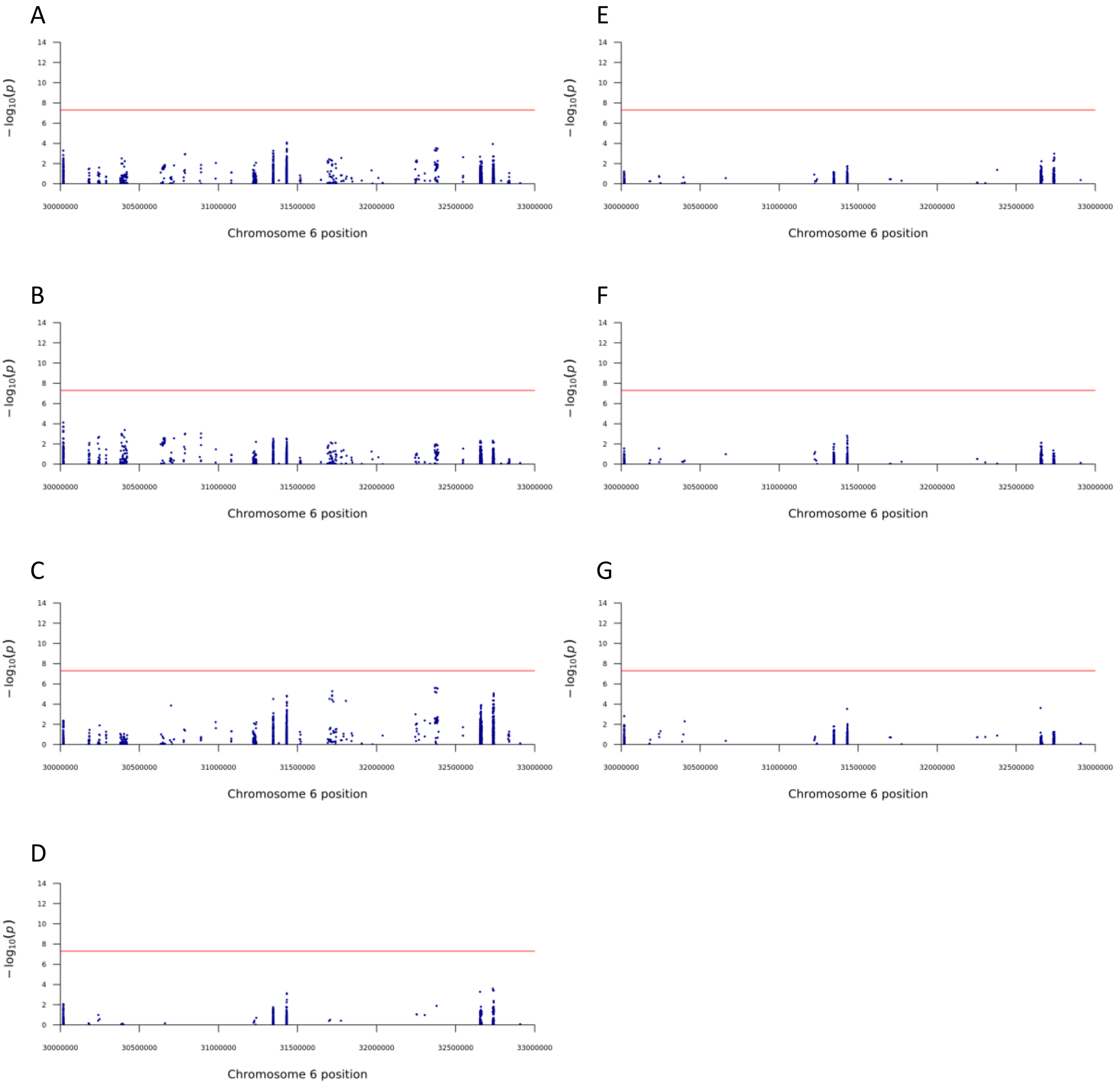


*Supplementary figure 3. Manhattan plots of HLA analysis for A) All Epilepsy, B) Focal Epilepsy, C) GGE, D) JME, E) Focal lesion negative, F) Focal due to other lesion, G) Focal HS.*

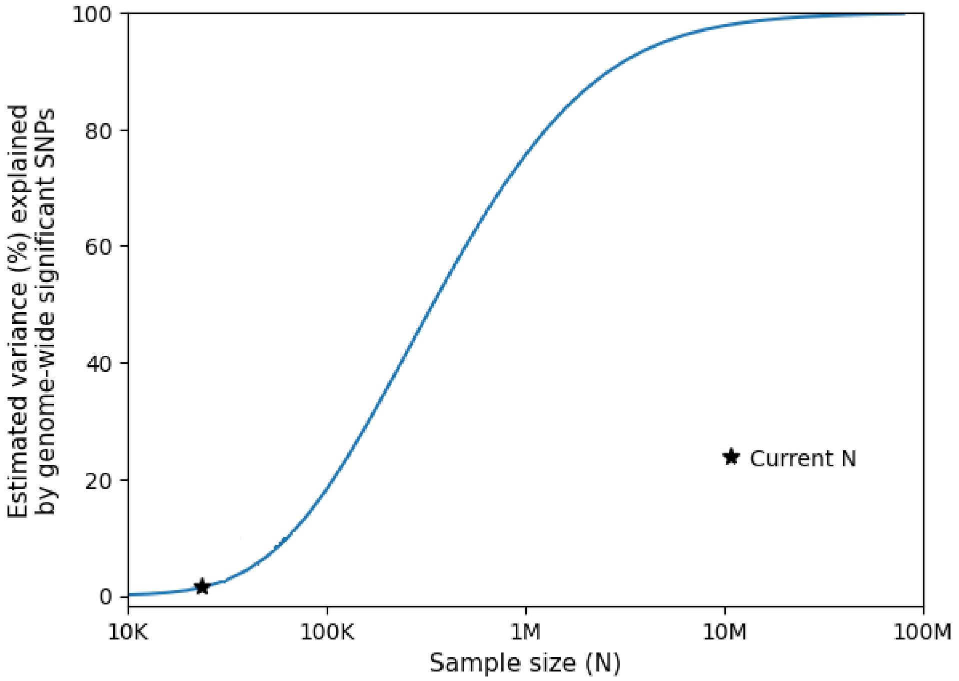


*Supplementary figure 4. Power analysis for GGE, using the MiXeR causal mixture model.[20] The X-axis shows the current and required sample size, and the Y-axis shows the corresponding explained variance by genome-wide significant SNPs at these sample sizes. An explained variance of 100% corresponds to the identification of all SNPs that underlie GGE SNP-based heritability.*

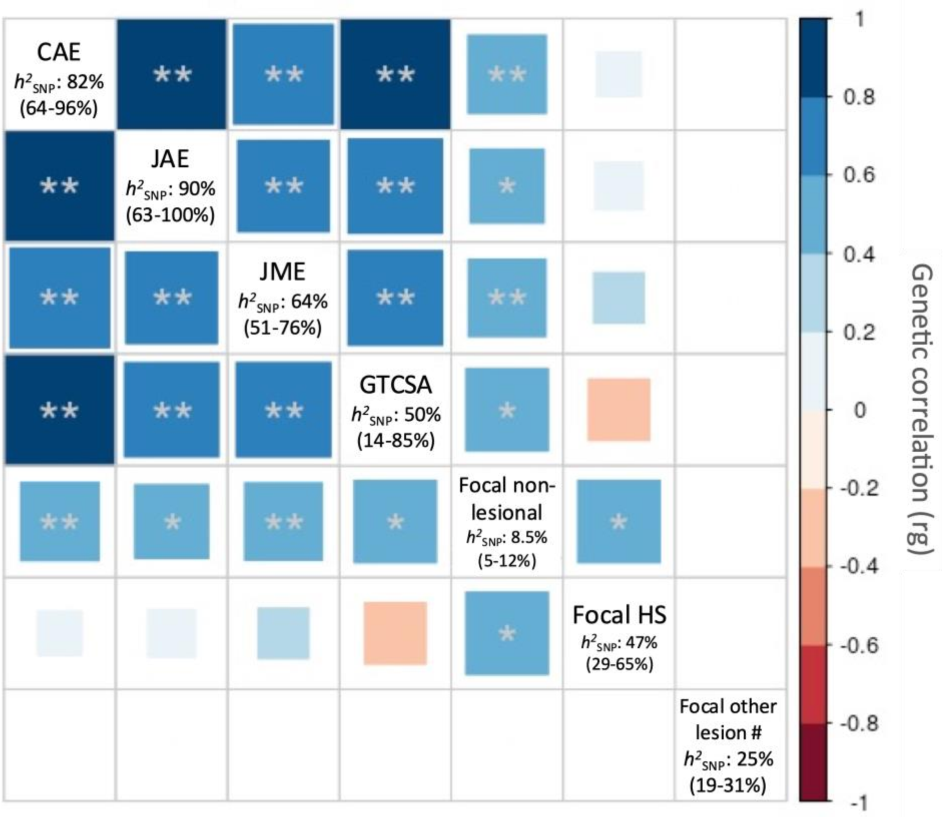


*Supplementary figure 5. Heritability estimates and genetic correlations between epilepsy syndromes. The genetic correlation coefficient was calculated with LDSC and is denoted by color scale from -1 (red) to +1 (blue). # rg out of bounds due to phenotype not reaching significant heritability; * P < 0.05, ** P < 0.0024 (Bonferroni correction).*

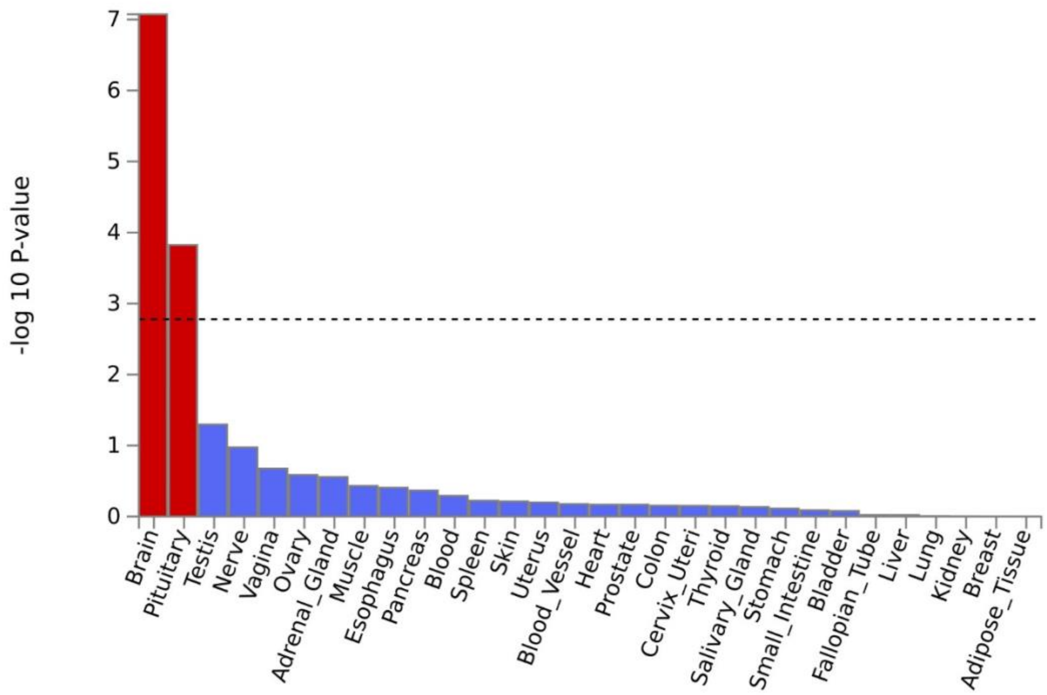


*Supplementary figure 6. Tissue-type enrichment of broad tissue types, as calculated with MAGMA,[21] using data from the Gene-Tissue Expression consortium (GTEx). The dotted line represents the significance threshold.*

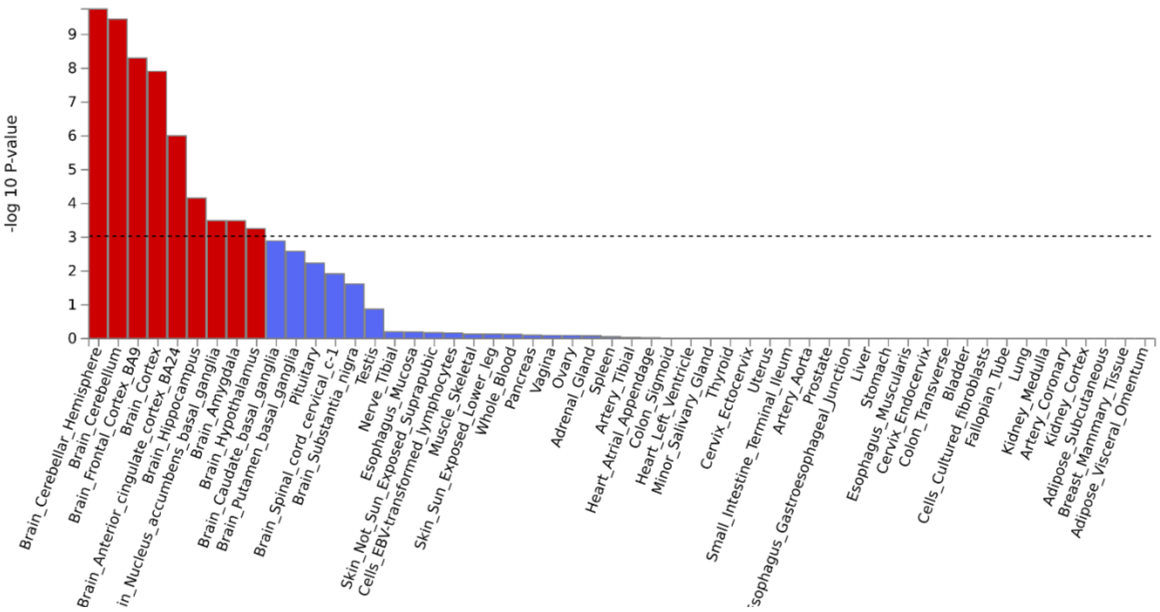


*Supplementary figure 7. Tissue-type enrichment of 54 tissues, including specific brain regions, as calculated with MAGMA[21] using data from GTEx. The dotted line represents the significance threshold.*

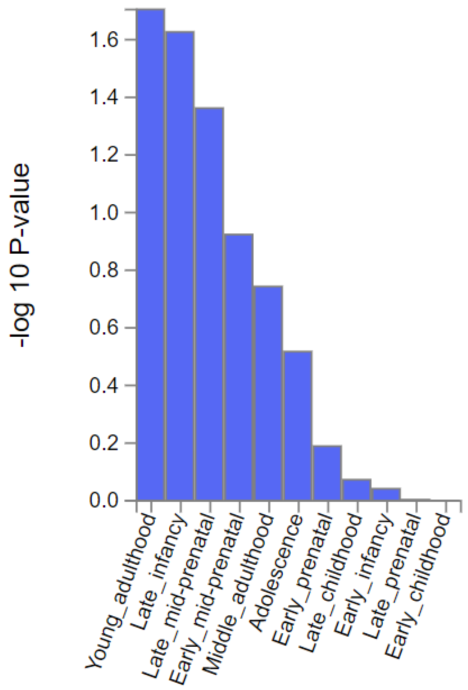


*Supplementary figure 8. Enrichment of genes expressed in the brain at 11 general developmental stages, as calculated with MAGMA,[21] using data from the BrainSpan consortium. The dotted line represents the significance threshold.*

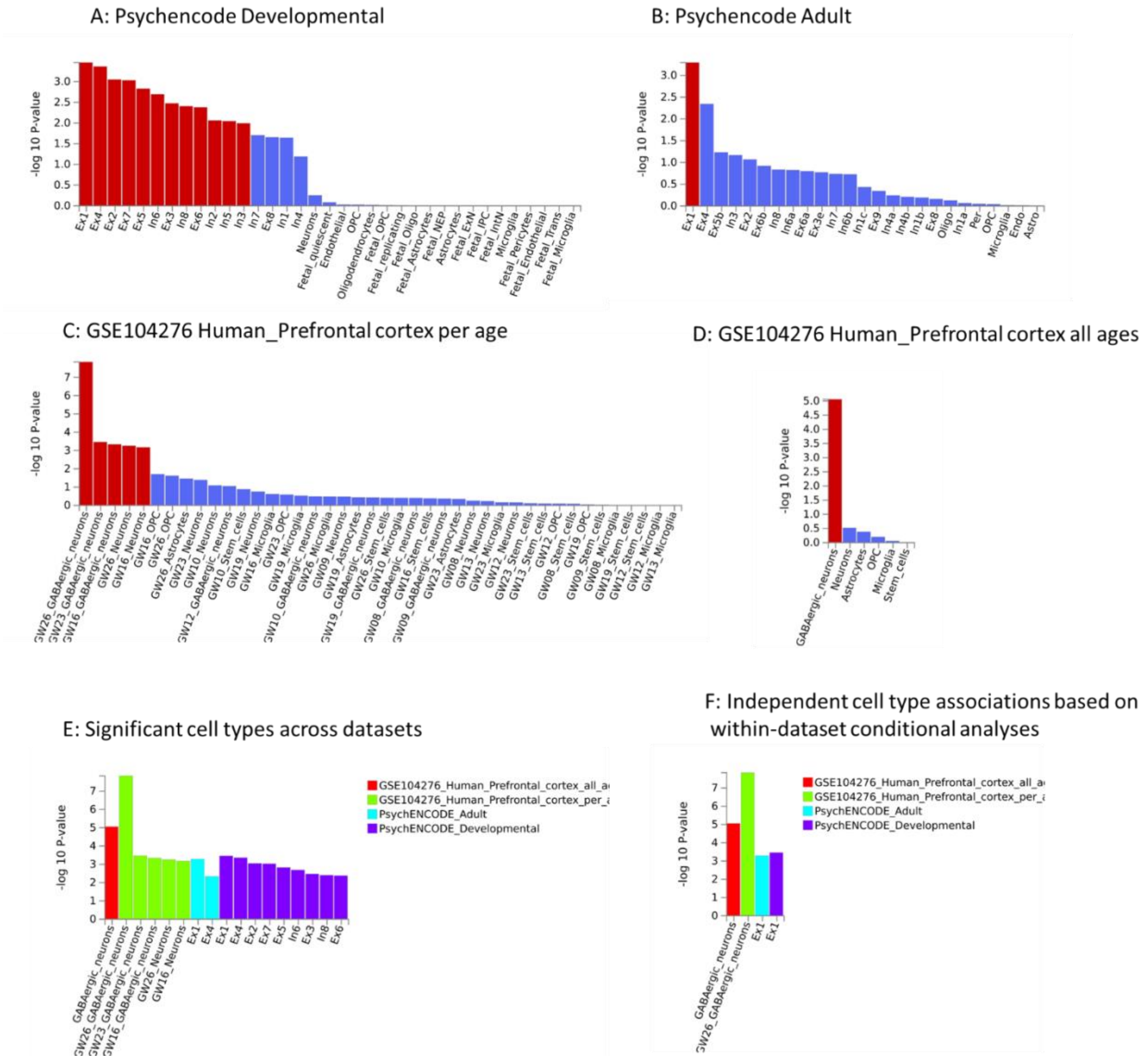


*Supplementary figure 9. Cell-type enrichment analyses across datasets, as calculated with FUMA.[22] Two different single-cell RNA sequencing datasets of human adult and developmental brain cells were assessed. Results from individual datasets are displayed in A-D with significant associations (after FDR correction) in red. Significant cell types across datasets are displayed in E, and significant cell-types after within dataset conditional analyses are displayed in F. Ex: excitatory neuron; In: inhibitory neuron.*

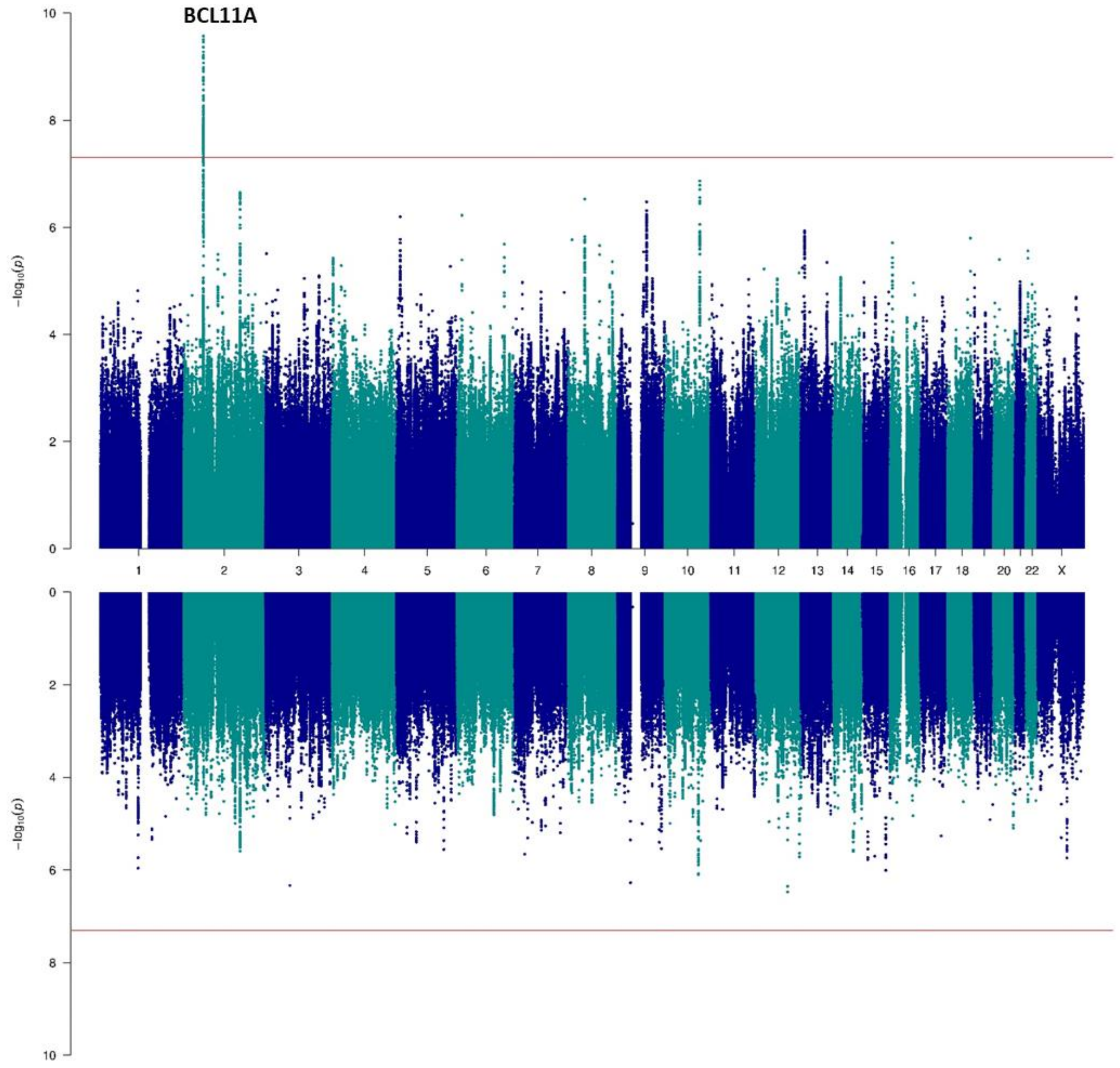


*Supplementary figure 10. Sex-specific GWAS of all epilepsy. The female-only is displayed at the top (n=13889 cases and 19676 controls) and male-only GWAS is displayed at the bottom (n=12259 cases and 18645 controls). We annotated genes that were implicated by our gene prioritization analyses.*

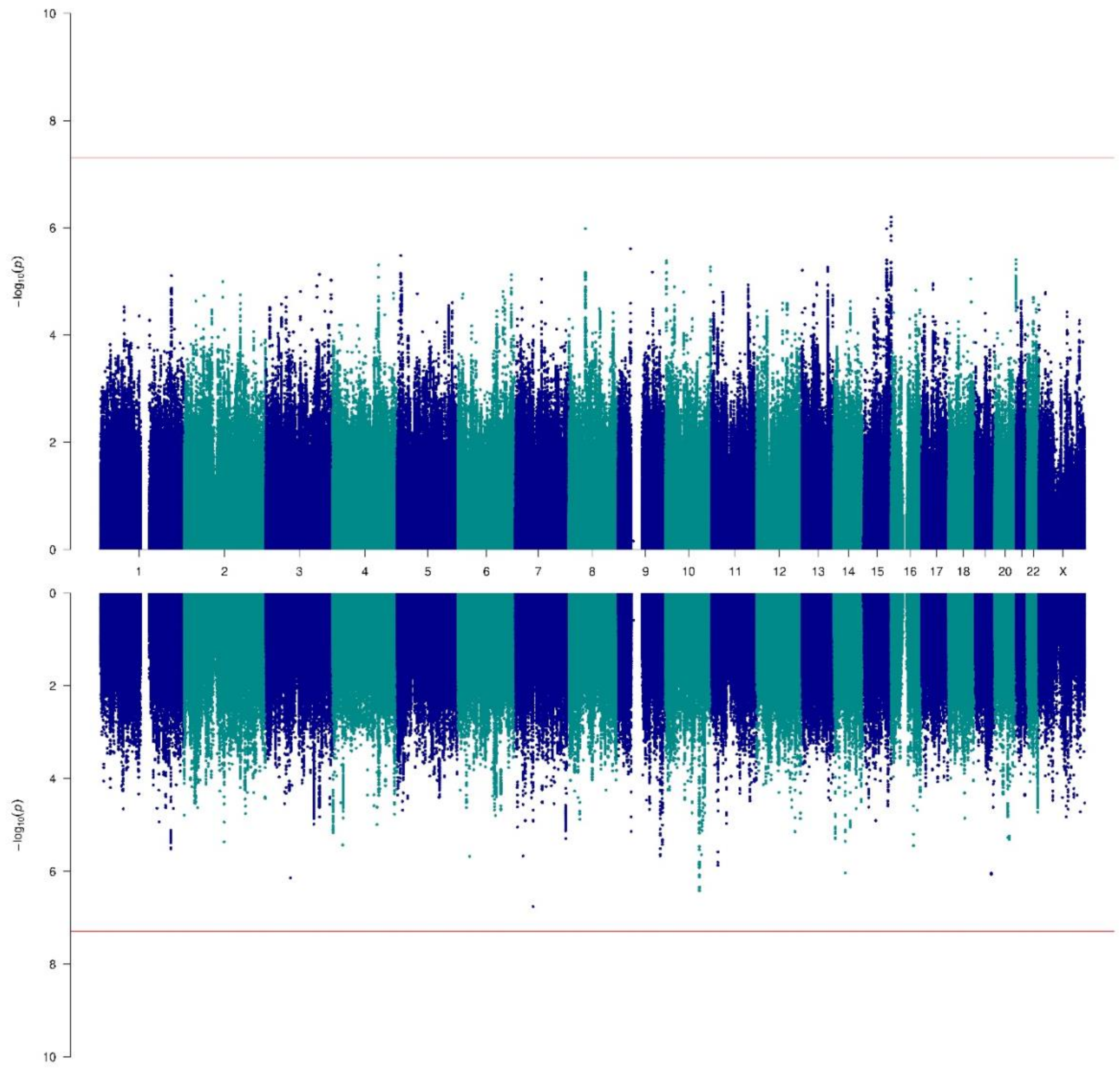


*Supplementary figure 11. Sex-specific GWAS of focal epilepsy. The female-only is displayed at the top (n=7175 cases and 19676 controls) and male-only GWAS is displayed at the bottom (n=6756 cases and 18645 controls).*

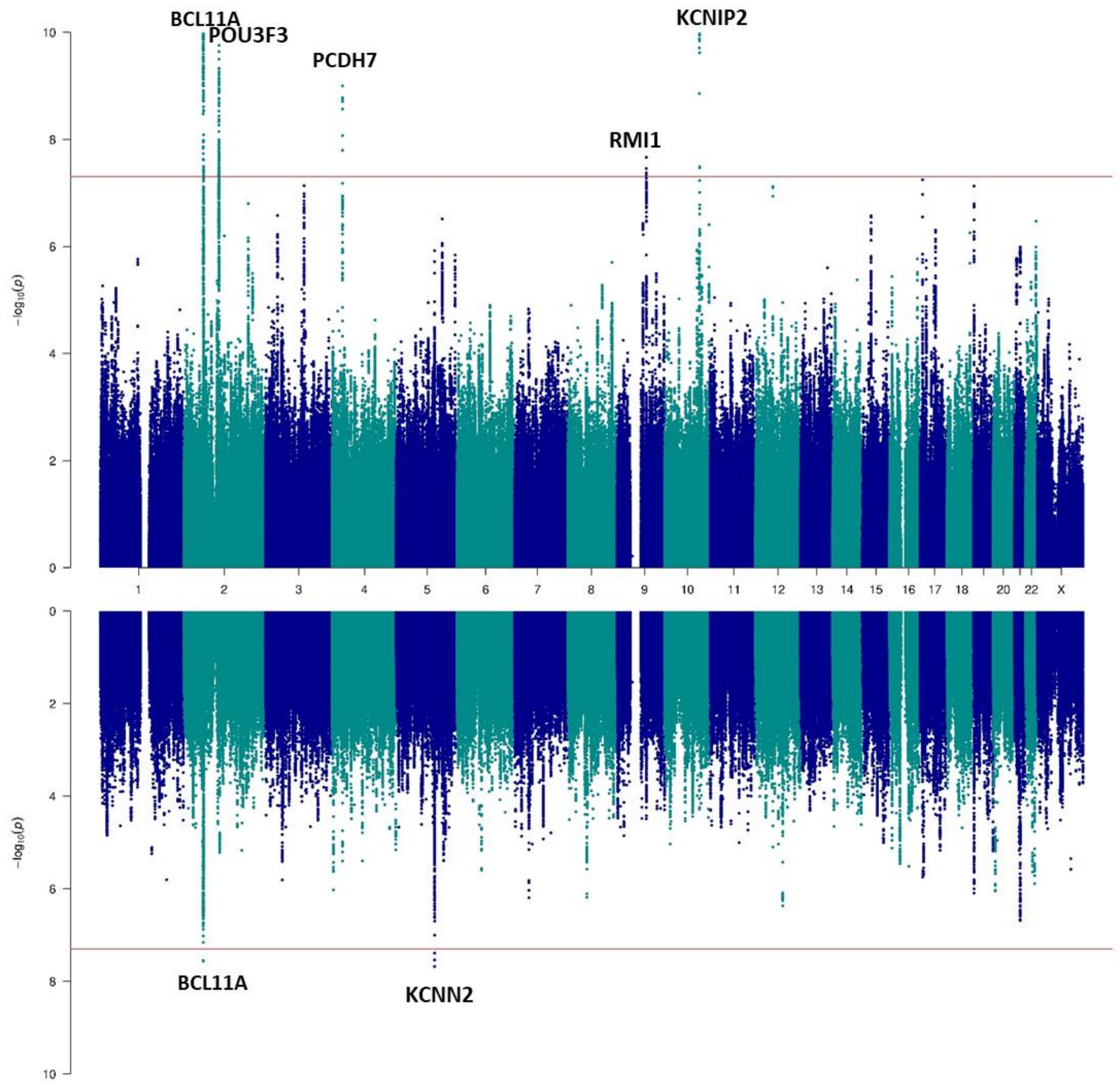


*Supplementary figure 12. Sex-specific GWAS of GGE. The female-only is displayed at the top (n=3946 cases and 19676 controls) and male-only GWAS is displayed at the bottom (n=2603 cases and 18645 controls). We annotated genes that were implicated by our gene prioritization analyses.*

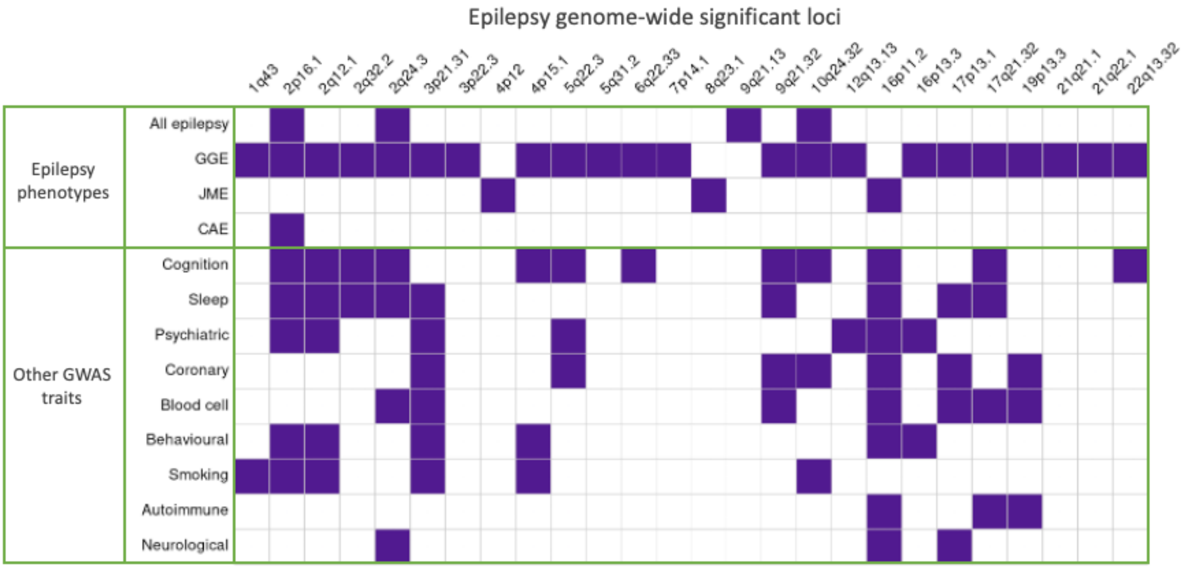


*Supplementary figure 13. GWAS traits each of the epilepsy genome-wide significant loci have been associated with indicated by a purple cell. Prior trait associations were determined by a p < 5*10-8 GWAS Catalog entry for the same SNP, or SNPs in high LD, as those reported in the epilepsy analysis.*

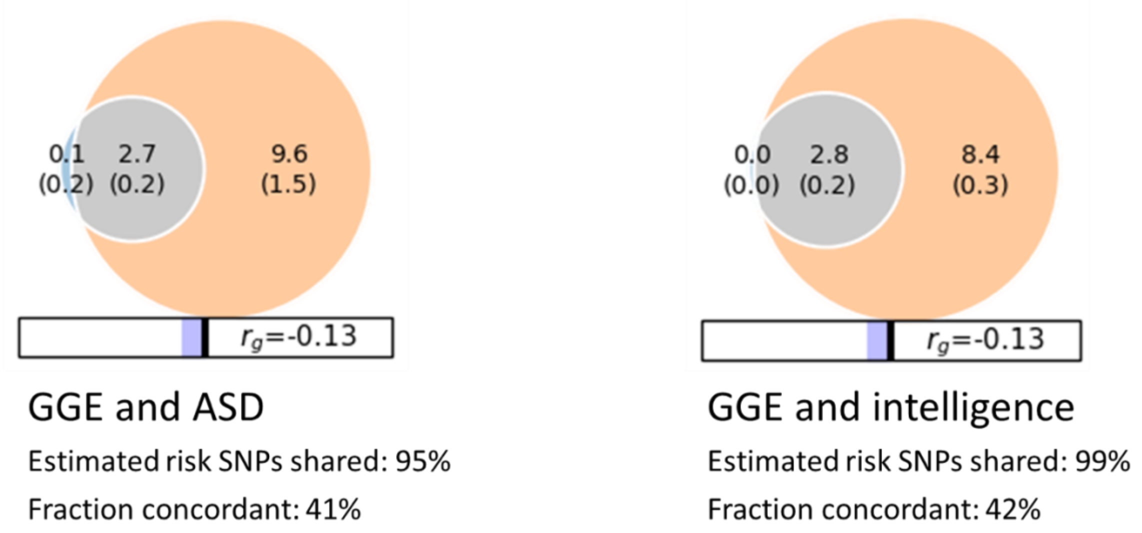


*Supplementary figure 14. Bivariate MiXeR analyses[20] showing the fraction of causal SNPs that are unique to GGE (blue), and shared (grey) and unique to ASD (left) or intelligence (right). Rg: genetic correlation coefficient.*

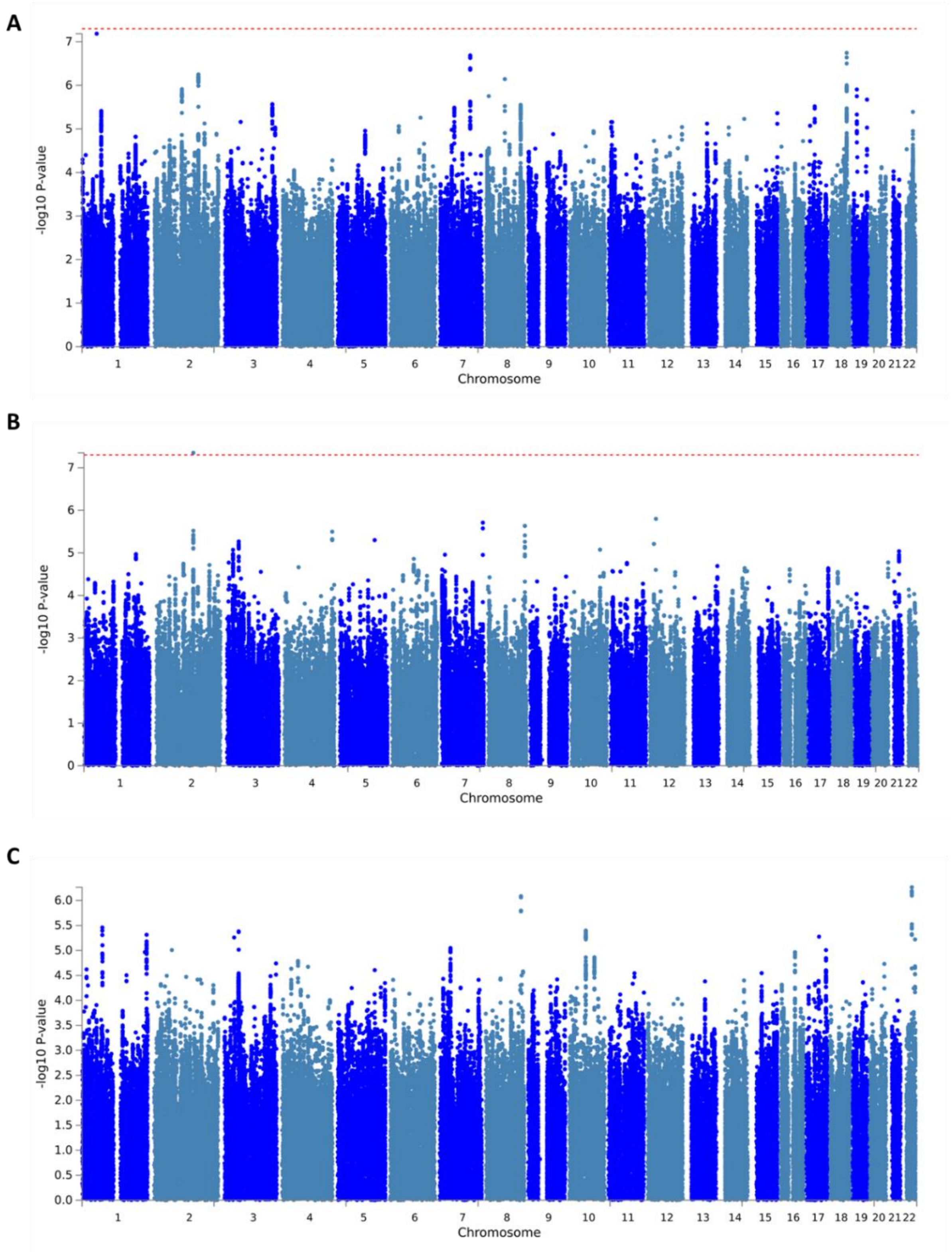


*Supplementary figure 15. Manhattan plots of Biobank-only GWAS of all (A), focal (B) and GGE (C). Chromosomal position is plotted on the X-axis and -log10 transformed P-values are plotted on the Y-axis.*

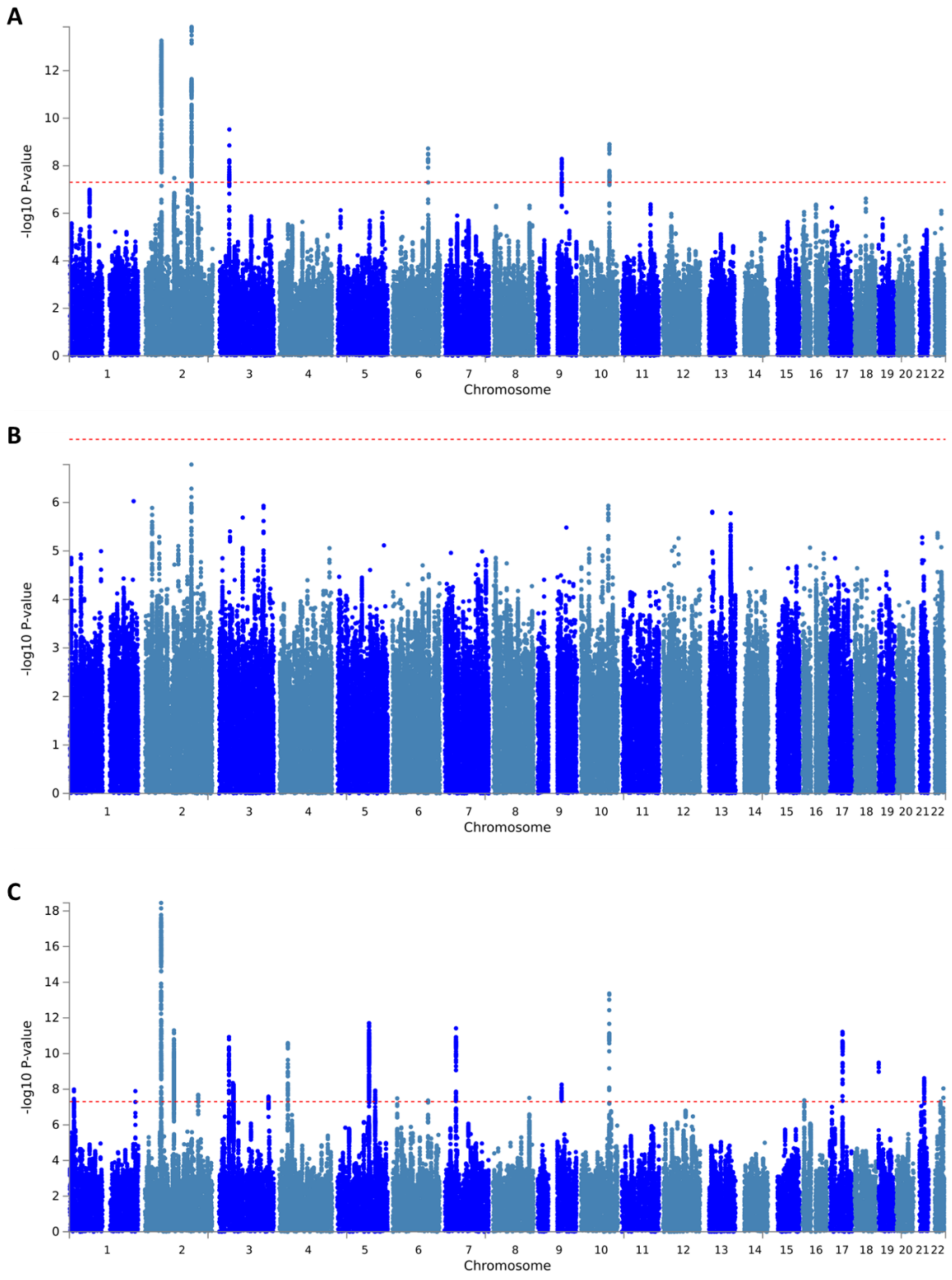


*Supplementary figure 16. Manhattan plots of meta-analysis combining the Biobanks with our primary GWAS of all (A), focal (B) and GGE (C). Chromosomal position is plotted on the X-axis and -log10 transformed P-values are plotted on the Y-axis.*

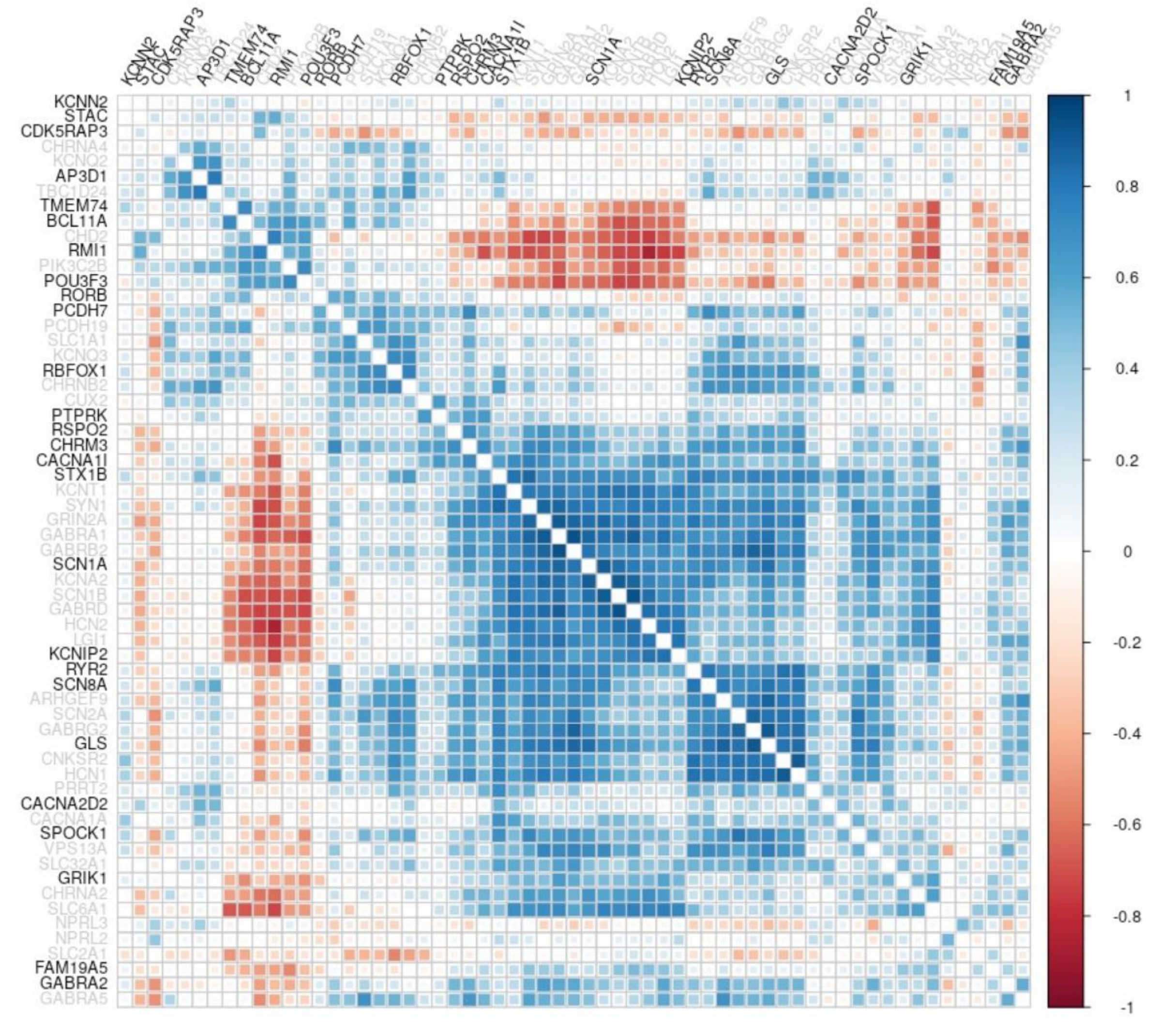


*Supplementary figure 17. Gene co-expression matrix produced by brain-coX[23] for known (grey) and candidate (black) epilepsy genes.*
